# Symptom-conditioned prediction of comorbidity in the hEDS–POTS–MCAS triad

**DOI:** 10.64898/2026.07.27.26358921

**Authors:** Advay Isaiah Banerji, Kyriakos Papas

## Abstract

**Background:** Hypermobile Ehlers–Danlos syndrome (hEDS), postural orthostatic tachycardia syndrome (POTS) and mast cell activation syndrome (MCAS) are reported to co-occur frequently. It is unclear how strongly, whether symptom profiles sharpen prediction of a second diagnosis given a first, and whether the published literature can support such inference at all.

**Methods:** We pooled 22 published cohorts—aggregate prevalence data, no primary human-subjects data—using Bayesian hierarchical random-effects models on the logit scale, and propagated the resulting posteriors through naive and tempered symptom updating. We introduce a feasibility screen derived from the Fréchet–Hoeffding bounds that tests whether separately pooled marginals can describe a single population, and we characterise the identifiability of latent class structure under disease-selected sampling.

**Results:** Directed comorbidity is strongly asymmetric: *π*_POTS|hEDS_ = 46.6% (95% CrI [32.5, 61.5]) against *π*_hEDS|POTS_ = 12.1% ([3.5, 38.5]), a near-fourfold gap, with *π*_MCAS|POTS_ lowest at 3.9% ([0.7, 21.6]). Prediction intervals exceed credible intervals throughout, indicating substantial between-cohort heterogeneity. The feasibility screen finds 26 of 102 testable cells (25.5%) incompatible with *any* joint distribution; critically, 21 of these fail the upper Fréchet bound and are invisible to the one-sided screen that is the natural first implementation. Among cells surviving the screen, symptom evidence is informative in four of six directions—ℙ(hEDS|POTS, **S**) rises from 12.1% to 49.7% on a four-symptom panel under tempered updating, and ℙ(POTS|MCAS, **S**) from 49.5% to 82.1%—but inert in both hEDS-cohort directions. A pathway-dispersion contrast excludes zero in two of six directions, in opposite signs and by margins of 0.1–0.2 percentage points, consistent with chance at this number of comparisons. We show latent class structure is not identified from disease-selected aggregate data, and that the single cohort reporting trivariate structure (*N* = 8) yields an exactly balanced table (OR = 1.00, 95% CI [0.063, 15.99]).

**Conclusions:** The pooled directional probabilities are usable as clinical priors, with intervals wide enough to preclude precision. Symptom-conditioned prediction is supported in some directions but not those most often invoked clinically, and every estimate rests on cohorts dominated by self-reported ascertainment. The principal methodological contribution is the two-sided feasibility screen: applied here it shows that a quarter of the testable literature cannot describe one coherent population, and that a one-sided implementation understates this sixfold.

## 1 Introduction

Hypermobile Ehlers-Danlos syndrome (hEDS), postural orthostatic tachycardia syndrome (POTS), and mast cell activation syndrome (MCAS) are chronic multisystem disorders that have been reported to overlap very frequently in patients. The Ehlers-Danlos syndromes (EDS) are heritable connective tissue disorders, and the most common type of EDS is the Hypermobile Ehlers-Danlos syndrome (hEDS), which is characterised by generalized joint hypermobility, musculoskeletal pain, and tissue fragility (Gensemer et al., 2020). Postural orthostatic tachycardia syndrome (POTS) is a common type of autonomic dysregulation associated with excessive heart rate when standing up (Zhao & Tran, 2023). The Mast Cell Activation Syndrome (MCAS) is a condition where mast cells are activated abnormally in an uncontrolled manner that often causes manifestations across different organs of the body (Özdemir et al., 2024). While these conditions differ in their pathophysiological origin, recent clinical evidence suggests substantial overlap in patient populations, with several authors proposing the existence of a frequently co-occurring clinical triad (Kohn & Chang, 2020). A paper published by Yao et al. (2025) has shown that 13% of 100 POTS patients appeared positive in hEDS using strict criteria and that 37% of them were diagnosed with MCAS using the consensus-2 criteria. Miller et al. (2020), Bishop et al. (2024), and Wallman et al. (2014) established similar results and raised attention to this trifecta.

Despite the increasing recognition that has been given to the hEDS-POTS-MCAS triad over the last decade, most of the existing literature focuses on investigating the potential biological mechanisms that link the conditions or describing the prevalence of symptoms between them. Fewer studies instead tried to evaluate how their overlapped symptom clusters could quantify the likelihood of a coexisting disorder. This could provide quantitative evidence regarding whether symptom patterns justify investigation for a coexisting disorder. It could also provide an established evidence-based association of symptom profiles and their comorbid diagnoses to assist clinical decision-making and help identify whether patients may benefit from further monitoring of these conditions without unnecessary investigations for cases that appear lower-risk.

This study specifically uses secondary clinical data and develops a quantitative Bayesian framework to evaluate whether symptom patterns can meaningfully alter the probability of a second diagnosis within the hEDS-POTS-MCAS triad. The research question therefore is: “To what extent can symptom patterns in patients with hEDS, POTS, and MCAS be used to estimate the likelihood of comorbid disease within the hEDS–POTS–MCAS triad?”.

## 2 Biological Background

### 2.1 Biological mechanisms underlying the hEDS–POTS–MCAS triad

Hypermobile Ehlers-Danlos syndrome (hEDS), postural orthostatic tachycardia syndrome (POTS), and mast cell activation syndrome (MCAS) are distinct disorders and they have different underlying pathophysiological mechanisms. However, their clinical manifestations arise through the dysfunction of major biological systems including connective tissue integrity, autonomic nervous system regulation, and mast-cell activation, and so all three are characterised by multisystem clinical manifestations (Gensemer et al., 2020; Zhao & Tran, 2023; Özdemir et al., 2024; Cuenca-Gómez et al., 2026). This means that patients affected by them are rarely only presenting one single symptom, but usually different clusters of syndromes that may even affect different organ systems. Clinicians therefore often identify the conditions through the symptom clusters that the patient presents (Castori, 2012; Kohn & Chang, 2020).

Some of the manifestations may reflect a biological mechanism caused by only one of the conditions, such as generalized joint hypermobility, which predominantly reflects connective tissue dysfunction and therefore usually only the presence of hEDS (Kohn & Chang, 2020; Monaco et al., 2022; Yao et al., 2025). Other manifestations -such as orthostatic intolerance or gastrointestinal dysmotility or chronic fatigue- can arise through different combined biological mechanisms and therefore occur in more than one of the conditions (Kohn & Chang, 2020; Monaco et al., 2022; Yao et al., 2025). The clinical manifestations observed in the hEDS-POTS-MCAS triad should be categorised into different symptom groups to distinguish between their root biological mechanisms and determine how they might connect (Castori, 2012; Song et al., 2021; Zhao & Tran, 2023; Özdemir et al., 2024).

This categorization is important because it helps interpret their potential clinical significance easier. Some symptoms could be disease specific, while others could indicate convergent manifestations between different pathophysiological processes. Accordingly, the following sections will examine the principal biological pathways that contribute to the different symptom groups, in order to explain their statistical relationships in the subsequent analysis.

### 2.2 Connective tissue dysfunctions

Musculoskeletal manifestations primarily arise from abnormalities that affect the structure and function of connective tissues, and they represent the most characteristic clinical feature that patients with hEDS present (Gensemer et al., 2020; Castori, 2012). Connective tissues are crucial for providing mechanical support to structures such as ligaments and tendons, so a disruption of their integrity can compromise joint stability and their regular biomechanical function (Gensemer et al., 2020; Castori, 2012).

Connective tissue abnormalities can result in generalized joint hypermobility and ligamentous laxity that can then lead to recurrent joint instability, subluxations, soft tissue injury, and repetitive mechanical stress for patients (Wang et al., 2025; Gensemer et al., 2020; Castori, 2012). Evidence does not primarily connect these manifestations to POTS and MCAS, allowing this path to be considered hEDS specific within this triad. (Kohn & Chang, 2020).

A major clinical symptom is chronic musculoskeletal pain. It develops through mechanical injury and tissue damage, that activate nociceptive pathways within bones, joints, ligaments, tendons, and muscles. This initiates peripheral inflammatory responses and pain signalling to the central nervous system (Puntillo et al., 2021), providing a direct mechanical explanation for how persistent pain is caused in patients with hEDS (Castori, 2012). In addition to chronic musculoskeletal pain, connective tissue dysfunction can contribute to several other clinically important musculoskeletal manifestations. Persistent joint instability and impaired proprioception increase the mechanical demands placed on surrounding muscles, often resulting in reduced muscle strength and endurance rather than primary muscular pathology (Castori, 2012; Voermans et al., 2010; Sahin et al., 2008). This functional muscle weakness may cause a repeating cycle of instability, contributing to symptoms associated with physical deconditioning. Furthermore, a proportion of patients with hEDS with prolonged widespread musculoskeletal pain may also fulfil the clinical criteria for Fibromyalgia, reflecting a contribution of central processing and sensitisation and not just connective tissue abnormalities (Rombaut et al., 2015).

However, although connective tissue dysfunction mechanisms are specific to hEDS here, pain, fibromyalgia and muscle weakness themselves should not be regarded as disease-specific. Rather, they represent multifactorial symptoms whose occurrence may reflect the interaction of structural, inflammatory, and neurophysiological mechanisms (Castori, 2012; Puntillo et al., 2021). Consequently, while generalized joint hypermobility is highly characteristic of hEDS, these broader musculoskeletal manifestations are not exclusively attributable to connective tissue dysfunction and may arise through alternative biological mechanisms in POTS and MCAS (Castori, 2012). The subsequent statistical analysis therefore evaluates these variables both individually and in combination with symptoms originating from other biological systems to determine whether they provide meaningful evidence of comorbid disease.

### 2.3 Autonomic nervous system dysfunction

Autonomic nervous system dysfunction is the major biological pathway connected to the cardiovascular and orthostatic manifestations most commonly associated with Postural Orthostatic Tachycardia Syndrome (POTS) (Blitshteyn, 2021). The autonomic nervous system is crucial for maintaining cardiovascular homeostasis as it regulates involuntary physiological processes such as heart rate, vascular tone, blood pressure, and tissue perfusion (Zhao & Tran, 2023). In normal physiological conditions, standing up results in a redistribution of blood towards the lower extremities. For this reason, a transient reduction in venous returns is compensated by autonomic reflexes that prevent significant reductions in blood pressure and tissue perfusion by increasing peripheral vasoconstriction and maintain cerebral blood flow (Zhao & Tran, 2023).

Patients with autonomic dysfunction, however, struggle with effective venous return as the compensatory autonomic mechanisms do not function effectively, giving reduced peripheral vasoconstriction, abnormal venous pooling, hypovolemia, or excessive sympathetic activation. This would reduce stroke volume upon standing up, and the compensation of the reduced cardiac output would come through excessive increase in heart rate, producing the characteristic orthostatic tachycardia observed in POTS (Zhao & Tran, 2023; Yao et al., 2025). Furthermore, such episodes can also reduce cerebral perfusion and so autonomic dysfunction overall gives rise a characteristic group of clinical manifestations, which include: dizziness, orthostatic intolerance, palpitations, tachycardia, presyncope, syncope, hypotension, and tremor (Zhao & Tran, 2023). Some secondary manifestations such as fatigue and cognitive impairment may also often happen due to repeated episodes of inadequate tissue perfusion but these include additional biological pathways as well that will be discussed later (Costa et al., 2023)

Although these manifestations are commonly grouped together as symptoms of autonomic dysfunction, they reflect different physiological consequences of autonomic nervous system impairment and therefore they may differ in their diagnostic significance across different patients (Sánchez-Manso et al., 2023). Palpitations and tachycardia primarily result from compensatory increases in the heart rate and excessive sympathetic activation, which occur when autonomic regulation fails to maintain an adequate cardiac output (Zhao & Tran, 2023; Yao et al., 2025). In contrast, dizziness, presyncope, and syncope develop when autonomic dysfunction causes insufficient cardiovascular compensation or transient reductions in blood pressure that produces inadequate cerebral perfusion (Zhao & Tran, 2023; Sánchez-Manso et al., 2023). Hypotension similarly reflects impaired autonomic control of vascular tone or abnormal vasodilation, and this makes it particularly relevant when considering autonomic dysfunction secondary to mast-cell mediator release in MCAS (Özdemir et al., 2024; Monaco et al., 2022; Sánchez-Manso et al., 2023). Finally, tremor is thought to arise predominantly from excessive sympathetic activity rather than impaired connective tissue integrity, and so it is more characteristic to hyperadrenergic autonomic states (Blitshteyn, 2021; Sánchez-Manso et al., 2023). These physiological differences suggest that autonomic manifestations should not be interpreted as a single uniform symptom group. Instead, analysing each manifestation individually may provide greater insight into the biological mechanisms underlying comorbid disease within the hEDS–POTS–MCAS triad.While autonomic dysfunction is regarded as the main biological explanation behind the cardiovascular manifestations characteristic to POTS, it should not be considered an exclusive indicator of it. In the case of hEDS, secondary autonomic dysfunction may arise as connective tissue abnormalities may often increase vascular compliance and promote venous pooling. This thereby predisposes affected individuals with hEDS to symptoms associate more closely to autonomic dysfunction as a secondary pathology originating from the primary pathology of the connective tissues (Kohn & Chang, 2020; Song et al., 2021; Cuenca-Gómez et al., 2026). Likewise, in MCAS, the abnormal activation of mast cells may influence autonomic regulation through the release of vasoactive mediators including histamine, prostaglandins, and other inflammatory molecules that alter vascular tone and cardiovascular responses (Özdemir et al., 2024; Monaco et al., 2022; Theoharides et al., 2024). Consequently, similar autonomic symptom profiles may arise through distinct biological mechanisms across all three disorders of the triad (Theoharides et al., 2024)

Therefore, symptoms associated with autonomic dysfunction, including dizziness/orthostatic intolerance, palpitations/tachycardia, syncope, presyncope, hypotension, and tremor will be analysed as variables that primarily occur to POTS patients and may co-occur to hEDS and MCAS patients too.

### 2.4 Mast-cell activation and inflammatory dysfunction

Mast-cell activation represents the principal biological pathway that underlies the allergic, inflammatory, and vasoactive manifestations observed within the hEDS–POTS–MCAS triad. Mast cells are tissue-resident immune cells that are distributed throughout the body, in connective tissues, the skin, gastrointestinal tract, respiratory system, peripheral nerves, and the cardiovascular system, where they participate in innate immune defence and they regulate vascular homeostasis and inflammatory responses. In physiological conditions, mast-cell activation occurs when exposed to pathogens or allergens and it results in the controlled release of inflammatory mediators of histamine, prostaglandins, leukotrienes, cytokines, and proteases. Collectively, these mediators regulate vascular permeability, smooth-muscle activity, immune-cell recruitment, and tissue repair (Özdemir et al., 2024; Monaco et al., 2022; Galván-Morales et al., 2025; Theoharides et al., 2024).

When this regulatory process becomes excessive or dysregulated, however, this persistent release of the mediators can simultaneously influence multiple organ systems negatively since mast cells are widely distributed throughout the body. Therefore, mast-cell activation gives rise to a phenotype of inflammatory symptoms that includes cutaneous, gastrointestinal, cardiovascular, respiratory, and neurological manifestations. While this biological pathway represents the defining pathological mechanism of MCAS, inflammatory mediator release may also contribute to symptom development in patients with hEDS and POTS without necessarily fulfilling the diagnostic criteria for MCAS (Kohn & Chang, 2020; Monaco et al., 2022; Cuenca-Gómez et al., 2026).

Although these manifestations are commonly considered together as consequences of mast-cell activation, their clinical presentation depends largely on the location and extent of inflammatory mediator release, and therefore they represent different inflammatory phenotypes. Distinguishing these manifestations is important for the present investigation because individual inflammatory symptoms may show different statistical associations with comorbid disease despite sharing the same biological pathway.

Within the skin, histamine-induced vasodilation together with increased vascular permeability primarily produces flushing and erythema, while activation of cutaneous sensory nerves gives rise to pruritus, itching, urticaria, and other hypersensitivity manifestations (Özdemir et al., 2024). When mediator release becomes more systemic, however, the same vasoactive mechanisms may produce widespread vasodilation and cardiovascular instability, contributing to hypotension and, in severe cases, anaphylaxis (Özdemir et al., 2024; Galván-Morales et al., 2025).

Because mast cells are also abundant throughout the gastrointestinal tract, inflammatory mediators released locally may alter smooth-muscle activity, increase epithelial permeability, stimulate intestinal secretion, and enhance visceral sensory signalling, thereby contributing to abdominal pain, nausea, bloating, gastroesophageal reflux, constipation, diarrhoea, and symptoms consistent with irritable bowel syndrome (Thwaites et al., 2022; Choudhary et al., 2021). Likewise, mast-cell mediators released around blood vessels may influence vascular tone and autonomic regulation, producing dizziness, tachycardia, orthostatic intolerance, and hypotension. This creates a clinical overlap with manifestations that are more commonly attributed to autonomic dysfunction (Monaco et al., 2022; Kohn & Chang, 2020).

Furthermore, inflammatory mediators released from activated mast cells can often influence connective tissue homeostasis, vascular regulation, autonomic function, peripheral nerve signalling, and gastrointestinal physiology (Theoharides et al., 2024). Consequently, mast-cell activation may contribute to symptom groups that are traditionally associated with connective tissue dysfunction or autonomic dysregulation despite arising through different primary pathological processes. Therefore, inflammatory manifestations frequently coexist with connective tissue and autonomic abnormalities, which makes it difficult to determine the underlying pathological process from individual symptoms alone (Monaco et al., 2022; Kohn & Chang, 2020; Cuenca-Gómez et al., 2026). Because this biological pathway extends across several organ systems these inflammatory manifestations will be evaluated both individually and in combination with other pathways.

### 2.5 Gastrointestinal dysfunction

Gastrointestinal dysfunction represents one of the most common cases where the triad’s conditions seem to converge in terms of their biological cause and their phenotype. Unlike the biological mechanisms discussed previously, gastrointestinal manifestations cannot be generally attributed to one pathological process, but simultaneously on many systems. A normal gastrointestinal physiology depends on connective tissue integrity, autonomic nervous system regulation, enteric neural signalling, smooth-muscle function, and local immune homeostasis. Therefore, a disruption of any of these biological systems can produce remarkably similar gastrointestinal symptoms despite the fact that they arise from fundamentally different pathological mechanisms (Aziz et al., 2025; Thwaites et al., 2022; Choudhary et al., 2021).

Motility-related manifestations, including constipation, diarrhoea, dysphagia, and bloating, primarily result from abnormalities in gastrointestinal transit, smooth-muscle function, or autonomic regulation (Aziz et al., 2025; Thwaites et al., 2022). Conversely, visceral sensory manifestations, including abdominal pain, nausea, dyspepsia, and symptoms consistent with irritable bowel syndrome, more commonly arise from visceral hypersensitivity, neuroimmune interactions, epithelial dysfunction, and inflammatory mediator release found mostly (Choudhary et al., 2021; Özdemir et al., 2024; Thwaites et al., 2022). Gastroesophageal reflux disease (GERD) may similarly result from impaired oesophageal motility, connective tissue laxity, autonomic dysfunction, or altered smooth-muscle regulation, illustrating how a single clinical manifestation may develop through multiple biological pathways (Thwaites et al., 2022; Aziz et al., 2025).

Consequently, interpreting how gastrointestinal symptoms overlap and what that says requires consideration of both their biological origin and the presence of accompanying manifestations from other physiological systems as similar symptom profiles may arise through distinct pathological mechanisms.

### 2.6 Neurovascular dysfunction and central nervous system manifestations

Another major example of biological convergence within the hEDS–POTS–MCAS triad is neurological dysfunction. Neurological manifestations frequently arise through the interaction of autonomic dysregulation, inflammatory mediator release, chronic pain, and impaired cerebral perfusion, and so the fact that they arise from such combinations rather than a single pathological mechanism, which makes it possible for symptoms affecting cognitive function and neurological performance to occur across all three disorders despite their different primary biological origins (Zhao & Tran, 2023; Özdemir et al., 2024; Cuenca-Gómez et al., 2026). The central nervous system depends on continuous cerebral perfusion, effective autonomic regulation, normal neuroimmune homeostasis, and balanced neuronal signalling in order to maintain cognitive performance, sensory processing, and normal neurological function. A disruption of any of these physiological systems may impair neuronal activity either directly through inflammatory signalling or indirectly through reduced cerebral blood flow and prolonged physiological stress (Blitshteyn, 2021; Theoharides et al., 2024; Sánchez-Manso et al., 2023).

Many factors in hEDS patients, such as persistent musculoskeletal pain, secondary autonomic dysfunction, and prolonged physical deconditioning, can collectively contribute to central sensitisation and chronic neurological symptoms, while connective tissue abnormalities remain the primary pathology (Castori, 2012; Rombaut et al., 2015; Cuenca-Gómez et al., 2026). Within patients with POTS, autonomic dysfunction frequently results in transient reductions in cerebral perfusion during orthostatic stress, which reduces the oxygen and nutrient delivery to the brain and contributes to symptoms including fatigue, cognitive impairment, dizziness, and headache (Zhao & Tran, 2023; Blitshteyn, 2021). In contrast, mast-cell activation can have an effect on neurological function through the release of inflammatory mediators that interact directly with systems such as peripheral nerves or autonomic ganglia, which promotes neuroinflammation, altered neuronal signalling, and neurovascular dysfunction (Theoharides et al., 2024; Özdemir et al., 2024).

Fatigue caused here primarily reflects impaired physiological homeostasis and reduced exercise tolerance that comes from autonomic dysfunction, chronic inflammation, prolonged pain, or combinations of these mechanisms (Zhao & Tran, 2023; Özdemir et al., 2024). Brain fog and other cognitive manifestations are more closely associated with impaired cerebral perfusion, neuroinflammation, and altered neuronal signalling that results in reduced attention, memory, and executive function (Blitshteyn, 2021; Theoharides et al., 2024). At the same time, Migraine and headache are thought to arise through interactions between neurovascular instability, autonomic dysregulation, inflammatory mediator release, and central sensitisation rather than any single pathological process (Theoharides et al., 2024). Lastly, sleep disturbances frequently develop as secondary manifestations of chronic pain, autonomic instability, and persistent inflammatory activity, while simultaneously contributing to fatigue, impaired cognitive performance, and central sensitisation (Rombaut et al., 2015).

The considerable overlap between these neurovascular and neuroimmune mechanisms explains why neurological manifestations are frequently observed across the hEDS–POTS–MCAS triad despite how their primary pathological origin differs. Consequently, to interpret how these manifestations co-exist requires consideration of both their underlying biological mechanisms and the presence of accompanying symptoms affecting other physiological systems (Cuenca-Gómez et al., 2026; Kohn & Chang, 2020; Theoharides et al., 2024).

### 2.7 Central sensitisation and psychosocial manifestations

The last biological pathway concerning the possible symptom profiles of the hEDS-POTS-MCAS triad that will be discussed in this investigation is central sensitisation and psychosocial manifestations. Central sensitisation refers to an increased responsiveness of the central nervous system to sensory stimuli, while psychosocial manifestations include symptoms affecting cognitive, emotional, and behavioural functioning such as anxiety, depression, sleep disturbance, chronic fatigue, and brain fog (Takeuchi et al., 2024; Woolf, 2011; Rombaut et al., 2015). These include downstream consequences of several other biological mechanisms already discussed and they are not just the result of one pathological process. For example, prolonged musculoskeletal pain, recurrent autonomic dysfunction, chronic inflammatory signalling, and repeated physiological stress may all contribute to widespread neurological symptoms, impaired sleep, fatigue, cognitive dysfunction, and altered emotional processing. (Rombaut et al., 2015; Castori, 2012; Takeuchi et al., 2024; Theoharides et al., 2024). Consequently, symptoms affecting psychological wellbeing are frequently observed across patients with hEDS, POTS, and MCAS despite the different primary pathophysiological mechanisms underlying each disorder. (Castori, 2012; Kohn & Chang, 2020; Cuenca-Gómez et al., 2026).

Two common symptoms within the triad the reflect exactly that are anxiety and depression are common symptoms. Takeuchi et al., 2024 explains that persistent autonomic instability, chronic inflammatory signalling, continuous nociceptive input, sleep disruption, and central sensitisation all influence neuronal processing within brain networks, regulating mood, cognition, and stress responses. However, anxiety more commonly reflects acute physiological hyperarousal resulting from excessive sympathetic activation, autonomic instability, and recurrent unpredictable symptom exacerbations. Depression, on the other hand, appears to develop more gradually through prolonged exposure to chronic pain, persistent fatigue, impaired physical functioning, sleep disturbance, and sustained neuroinflammatory signalling, representing the cumulative burden of long-term multisystem disease rather than transient physiological stress (Takeuchi et al., 2024; Berglund et al., 2015; Theoharides et al., 2024). Anxiety is therefore closer to autonomic dysfunction and physiological hyperarousal (Blitshteyn, 2021; Zhao & Tran, 2023), while depression more frequently reflects the cumulative burden of chronic multisystem illness rather than one isolated biological mechanism. (Berglund et al., 2015; Castori, 2012). Despite these differences, substantial biological overlap exists because chronic autonomic dysfunction, inflammation, persistent nociceptive signalling, and central sensitisation may all interact simultaneously, meaning that patients frequently present features of both manifestations.

Overall, anxiety and depression should therefore not be interpreted as manifestations that are specific to one disease of the triad, but as downstream clinical consequences that integrate the effects of multiple interacting biological pathways discussed throughout this background. This means that evaluating these manifestations alongside musculoskeletal, autonomic, inflammatory, gastrointestinal, and neurovascular symptoms may provide additional insight into the complex biological phenotype associated with comorbid disease within the hEDS–POTS–MCAS triad. (Kohn & Chang, 2020; Cuenca-Gómez et al., 2026).

## 3 Methods

### 3.1 Study design

This investigation employed a retrospective quantitative observational study design, with the use of secondary data extracted from published clinical studies, patient registries, cohort studies, and case series that investigate the conditions of the triad hEDS-POTS-MCAS and different variables affecting them. The aim of this was to create a unified symptom database for the statistical analysis through the integration of study-level and patient-level data reported in the literature, thereby allowing the symptom patterns to be examined across published populations that represent large cumulative numbers of patients.

The investigation was conducted using two main complementary analytical levels. Firstly, study-level prevalence data was used from published cohorts and registries that report a specific sample size for one primary diagnostic population and the prevalence of individual symptoms withing that population. In some cases, the occurrence of hEDS, POTS, or MCAS within that population was also reported which gave further information for the statistical analysis. These data were then used to compare symptom prevalence and broader symptom patterns across the three diagnostic populations. Secondly, detailed case-level information was also used, as it reports the occurrence of specific symptoms within individual patients and therefore allows to directly view within-patient symptom co-occurrence, something very meaningful statistically. However, such patient-level data were considerably less available in published literature found, and therefore this level of analysis was limited to a small sample of eight patients. These data were separated into a distinct patient-level dataset. The study-level and patient-level datasets were analysed separately where it was required, as aggregated prevalence values cannot be interpreted as complete individual patient profiles.

The primary aim of the investigation was aimed to be a proper evaluation of whether patterns of symptom presentation are associated with the occurrence of comorbid disease within the hEDS– POTS–MCAS triad, and to provide quantitative insight into how the observed likelihood of each condition differs across symptom profiles. The study therefore does not aim to establish a perfect diagnostic tool, but to investigate whether individual clinical manifestations and combinations of symptoms demonstrate statistically meaningful associations with the presence of one or more disorders within the triad. Where consistent statistical patterns are identified, information may be provided to develop a quantitative basis for determining which symptom profiles warrant further investigation in future clinical research.

Because the investigation combines information originating from studies with different populations, methodologies, and diagnostic criteria, the statistical analyses are interpreted as measures of association rather than direct estimates of disease risk. Biological mechanisms were also discussed in the preceding chapter as they are subsequently used to interpret the statistically identified symptom patterns and their consistency with current understanding of the pathophysiology underlying the three disorders.

### 3.2 Data sources and study selection

Published studies used were identified through searching scientific literature databases and academic search platforms, primarily PubMed and Google Scholar. Additionally, papers were also found through the reference lists of review articles concerning hEDS, POTS, MCAS, and their clinical overlap. Other studies that were needed for the theoretical background were identified when specific biological mechanisms or clinical manifestations required further supporting evidence for the interpretation of the quantitative data.

Studies were considered eligible for being entered in the study-level quantitative dataset when they investigated an identifiable population affected by fully or partly by at least one of hEDS, POTS, or MCAS and explicitly reported numerical prevalence or frequency data for at least one clinical manifestation included in the investigation. The study sample size and primary diagnostic population were also required to be identifiable so that the reported symptom values could be interpreted in relation to the appropriate disease population. Studies were not required to report every selected symptom, as the clinical variables assessed differed considerably between published investigations. Symptoms that were not assessed or numerically reported by an individual study were therefore treated as missing data rather than as absent manifestations. Due to the fact that the studies selected may have different origins and purposes, a source bias was always taken into consideration. For example, a study focused on MCAS patients with vomiting problems might have provided important data for other symptoms, but the GI symptoms variable is obviously biased, and this was aimed to be taken into consideration throughout the numerical analysis.

Studies were not included in the primary study-level prevalence dataset if no numerical symptom data was given inside. This means that any kind of study providing exclusively mechanistic or descriptive information, or consisted solely of diagnostic recommendations or clinical guidelines, was not included. Studies were also excluded from the dataset when they did not provide sufficient information to identify the population from which the reported values originated. Case reports and detailed case series were also not combined with the aggregated prevalence dataset because the symptom information reported in prevalence sheets cannot directly tell the co-occurrence of symptoms, and so that would be a wrongful comparison that would lead to inaccurate results. Where sufficiently detailed individual symptom profiles and diagnostic information were available, however, these data were considered separately for inclusion in the patient-level dataset described in Section 3.1.

A distinction was therefore maintained between literature used as a theoretical source and studies used for quantitative data extraction. Review articles, mechanistic studies, and diagnostic frameworks were useful in their use to establish the biological and clinical interpretation of the selected symptoms, but their qualitative claims were not converted into numerical prevalence estimates. Because the primary objective of the investigation was to compare symptom patterns across the hEDS–POTS–MCAS triad, study selection prioritised sources that provided information relevant to the predefined symptom variables or the occurrence of comorbid conditions within the triad. The final evidence base therefore incorporated studies with different sample sizes and observational methodologies. These differences were retained and subsequently considered during data processing and statistical interpretation.

## 4 Statistical analysis: aims and contributions

The biological account of Section 2 organizes the clinical manifestations of the triad by pathway and asks a quantitative question: to what extent do symptom patterns carry information about undiagnosed comorbidity? Tackling this question precisely requires separating it into four distinct problems, each in its own direction, which differ sharply in how well the available evidence can address them:

### (Q1) Regarding marginal comorbidity

Given that a patient carries a diagnosis *A*, what is the probability that the patient also carries diagnosis *B*? This is a question about six directed quantities, not three symmetric ones, since ℙ(*B* | *A*)*≠* ℙ(*A* | *B*) whenever the marginal prevalences differ.

### (Q2) Regarding symptom-conditioned prediction

Given some diagnosis *A* and an observed symptom profile, how should the probability of *B* be updated? This is a question about the informational content of symptoms over and above the base rate.

### (Q3) Regarding latent structure

Is the observed clustering better explained by three distinct disease processes that happen to co-occur, or by a smaller number of latent phenotypes of which the three diagnoses are partial manifestations?

### (Q4) Regarding monitoring decision

Combining (Q1) and (Q2): does a given symptom profile raise the posterior probability of a second diagnosis above a clinical threshold, at which further monitoring would be warranted? Also, (motivated by the biological organization of symptoms into pathway groups) do symptoms that are drawn from *multiple* biological systems provide more such evidence than an equal number drawn from a single system?

Questions (Q1)–(Q3) are progressively more ambitious and, as we shall establish, progressively less supported by the obtained evidence. (Q4) is the decision-theoretic synthesis that the clinical motivation demands. A central thesis of this paper is that these questions have sharply different *identifiability* status given aggregate data, and that conflating them produces analyses whose apparent rigor exceeds their evidential warrant.

### Contributions of the statistical analysis

1. A hierarchical Bayesian meta-analysis of all six directed comorbidity probabilities (Section 4.3.1), with weakly informative priors chosen specifically to avoid false precision in directions supported by as few as two studies.
2. A reference-group deconvolution identity (Lemma 4.3.2) that makes the symptom-conditioned likelihood ratio internally consistent with the comorbidity prior, replacing the makeshift denominator used in naive applications.
3. A proof that this identity carries a Fréchet-type feasibility constraint (Proposition 4.3.5), and the empirical finding that the constraint is violated for 47 out of 104 symptom–direction cells (a quantitative diagnostic of inconsistency in the published literature (Section 5.2)).
4. Two non-identifiability results for latent class analysis: a constructive counterexample from marginals (Theorem 4.3.14) and a structural degeneracy result for disease-selected cohorts (Proposition 4.3.18), together with positive verification that the estimator itself is correct (Section 5.5).
5. Exact recovery of a 2 *×* 2 joint contingency table from the single trivariate reporting cohort by inclusion–exclusion (Proposition 4.3.19), establishing that the corpus contains no individual-level joint information capable of constraining the triad, together with the documented correction of an earlier erroneous extraction that had appeared to show the opposite (Remark 5.6.2).
6. A pathway-structured evaluation of symptom evidence (Section 5.3) that operationalises the biological grouping of symptoms into connective-tissue, autonomic, mast-cell, gastrointestinal, neurovascular, and central-sensitisation systems, and tests whether cross-pathway symptom combinations carry more comorbidity evidence than within-pathway combinations of equal size; the test returns a null result in five of six directions, which we report as such.
7. A decision-analytic layer (Section 5.4) that maps posterior probabilities onto explicit monitoring thresholds, answering the clinical triage question (Q4) by identifying, per direction, the number and type of symptoms required to cross a given threshold.

### 4.1 Background and theoretical foundations

#### 4.1.1 Notation

##### Definition 4.1.1

(Diagnostic and symptom variables). Let *D* = {POTS, hEDS, MCAS} index the three diagnoses. For a patient drawn from a population of interest, let *D*_*A*_ ∈ {0, 1} denote the indicator of diagnosis *A* ∈ *D*. Let *S* = *{S*_1_, …, *S*_*J*_ } denote a finite list of symptoms, with *S*_*j*_ ∈ {0, 1} being the indicator that symptom *j* is present. We write *A* for the event *{D*_*A*_ = 1} and *¬A* for its complement *{D*_*A*_ = 0}, and abbreviate ℙ(*B* | *A*) for ℙ(*D*_*B*_ = 1 | *D*_*A*_ = 1).

##### Definition 4.1.2

(Directed comorbidity probability). For distinct *A, B* ∈ *D*, the *directed comorbidity probability* is *π*_*B*|*A*_ := ℙ(*B* | *A*). The six quantities *{π*_*B*|*A*_}_*A*≠*B*_ are the primary estimands of (Q1). We emphasize that *π*_*B*|*A*_ and *π*_*A*|*B*_ are distinct parameters related by Bayes’ theorem,

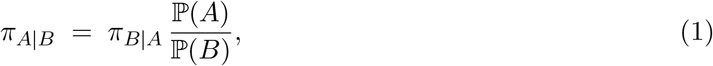

so that any observed asymmetry between them would, in the first instance, be attributable to differing marginal prevalences rather than to asymmetric association. We stress that this attribution cannot be verified here: by Proposition 4.3.18 the marginals ℙ(*A*) and ℙ(*B*) are not identified by disease-selected data, so (1) cannot be used to check the mutual consistency of our six estimates.

##### Definition 4.1.3

(Logit and expit). For *p* ∈ (0, 1) define logit(*p*) = log *p/*(1 −*p*) and its inverse expit(*θ*) = (1 + *e*^−*θ*^)^−1^.

##### Definition 4.1.4

(Odds and likelihood ratio). The odds of an event *E* are odds(*E*) = ℙ(*E*)*/*(1 − ℙ(*E*)). For a binary observation *S* and hypothesis *B* conditioned on background information *A*, the *likelihood ratio* is

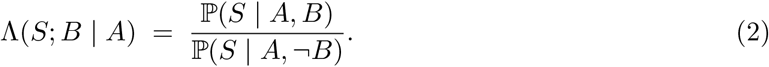

#### 4.1.2 Bayesian updating in odds form

##### Lemma 4.1.5

(Sequential odds update). *Let S*_1_, …, *S*_*n*_ *be binary observations. If the conditional independence condition*

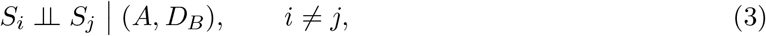

*holds, then*

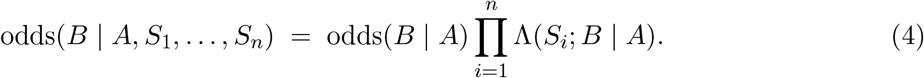

*Proof*. By Bayes’ theorem applied to the event *B* conditional on *A* and the vector **S** = (*S*_1_, …, *S*_*n*_),

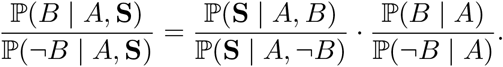

The normalizing constant ℙ(**S** | *A*) cancels between numerator and denominator, which is precisely the motivation for working in odds. Under (3) the joint conditional densities factorize, ℙ(**S** | *A, D*_*B*_) = Π_*i*_ ℙ(*S*_*i*_ | *A, D*_*B*_), and the ratio of products is the product of ratios, giving (4).

##### Remark 4.1.6

(The status of (3)). Condition (3) is the *naive Bayes* assumption. It is not a technical convenience but a substantive claim about symptom biology: it asserts that, among patients with fixed diagnostic status, the presence of fatigue conveys no information about the presence of cognitive dysfunction. This is implausible on mechanistic grounds. Section 4.3.2 introduces a tempered update that partially accommodates violation, and Section 7discusses why the assumption cannot be tested with the present data.

#### 4.1.3 Meta-analysis of proportions

##### Definition 4.1.7

(Random-effects model for proportions). Let studies *i* = 1, …, *k* report *y*_*i*_ events among *n*_*i*_ subjects. The random-effects binomial–logit–normal model posits

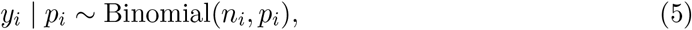

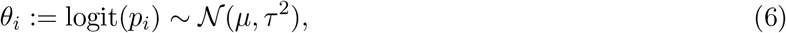

independently across *i*. The parameter *µ* is the pooled log-odds and *τ* ^2^ the between-study variance on the logit scale.

The model separates two sources of variation. Sampling variability is captured by (5); genuine heterogeneity in the underlying population parameter across study contexts is captured by (6). The fixed-effect model is the degenerate case *τ* = 0, which asserts that all studies estimate a common *p*. For symptom prevalences pooled across cohorts differing in ascertainment protocol, diagnostic criteria, referral pattern, and geography, *τ* = 0 is untenable a priori, and we do not entertain it. We report *τ* itself rather than the more familiar *I*^2^ of Higgins and Thompson (2002), which is derived from Cochran’s *Q* (1954): *I*^2^ is a ratio of variance components rather than an absolute measure, is poorly estimated at the study counts available here, and is not directly interpretable on the scale of the estimand.

##### Definition 4.1.8

(Pooled estimand and predictive distribution). The *pooled probability* is expit(*µ*). The *posterior predictive* probability for a new, previously unobserved study is expit(*θ*_new_) where *θ*_new_ ∼ *N* (*µ, τ* ^2^). The corresponding 95% intervals are termed the *credible interval* and the *prediction interval* respectively (Higgins et al., 2009; IntHout et al., 2016).

The credible interval quantifies uncertainty about the population-average parameter; the prediction interval quantifies where a future study’s parameter is expected to fall. When *τ* is large the two diverge sharply, and reporting only the former conveys a misleading impression of homogeneity. We report both throughout.

#### 4.1.4 The choice of prior on *τ*

With *k* as small as two, the between-study variance is very weakly identified by the likelihood. Method-of-moments estimators such as DerSimonian–Laird (1986) frequently return 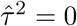 in this regime, collapsing the random-effects model to the fixed-effect model and producing intervals that are too narrow by a large factor. This is not a defect of a particular estimator but a generic consequence of attempting to estimate a variance component from a handful of observations.

We therefore adopt a weakly informative half-normal prior (Gelman, 2006; Röver et al., 2021; Turner et al., 2015),

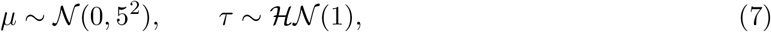

which places no mass at *τ* = 0 as a point atom, permits substantial heterogeneity, and regularizes away from the implausible tail *τ* ≫ 1. On the logit scale a value *τ* = 1 corresponds to a roughly threefold spread in odds between typical studies, which is a generous but not absurd allowance. The prior on *µ* is diffuse: *N* (0, 25) on the log-odds scale covers (10^−4^, 1 − 10^−4^) on the probability scale to within two standard deviations.

Because *τ* is weakly identified, inferences about *τ* itself are prior-sensitive by construction, and we make no strong claims about its numerical value. Inferences about expit(*µ*) are considerably more robust, since *µ* is informed directly by the study-level counts. The appropriate reading of a large posterior *τ* is qualitative—”these studies disagree more than sampling error explains”—rather than quantitative.

#### 4.1.5 Ecological inference and identifiability

The recovery of individual-level relationships from group-level aggregates is the classical problem of *ecological inference* (Robinson, 1950; Greenland & Robins, 1994), and the obstacle it presents here is one of identifiability rather than precision.

##### Definition 4.1.9

(Identifiability). A parameter *ϕ* of a statistical model *{P*_*ϕ*_ : *ϕ* ∈ Φ} is *identified* by an observation functional *T* if 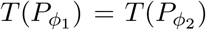 implies *ϕ*_1_ = *ϕ*_2_. If distinct parameter values induce identical observables, *ϕ* is *not identified*, and no estimator—however efficient, and regardless of sample size—can consistently recover it.

Definition 4.1.9 is the pivot of this paper. Non-identifiability is not remedied by larger *n*, it is a property of the map from parameters to observables. An estimation routine applied to a non-identified parameter will typically still *converge*, and will report an answer, but that answer is determined by the arbitrary features of the algorithm—initialization, penalization, stopping rule— rather than by the data. We exhibit exactly this pathology empirically in Section 5.5.

### 4.2 Data

#### 4.2.1 Provenance and structure

The primary corpus comprises 22 published studies. One row was entered without a study identifier and has since been traced to the claims-database cohort of Kozyra et al. (2024); it contributes three symptom prevalences, its diagnostic marginal being degenerate and therefore uninformative under Proposition 4.3.18. The corpus includes large POTS survey and clinic cohorts (Kohno et al., 2021; Shaw et al., 2019), EDS systemic-manifestation series (Yao et al., 2025), and the triad-focused cohort of Wang et al. (2021). Diagnoses follow the prevailing criteria of the source studies— the 2017 international classification for hEDS (Malfait et al., 2017) and consensus statements for POTS (Freeman et al., 2011; Sheldon et al., 2015)—a heterogeneity we revisit in Section 7. Each row records: a study type descriptor; a sample size *n*_*i*_; a *cohort type* indicating the diagnosis on which the cohort was selected; up to 29 symptom prevalence percentages; and up to four diagnostic prevalence percentages (POTS, hEDS, MCAS, and “all three”). A complete description of all included studies, together with the variables extracted from each source, is provided in the Appendix (Tables A1–A2).

##### Definition 4.2.1

(Cohort selection variable). Each study *i* is associated with a selection diagnosis *A*(*i*) ∈ *D* such that every subject in study *i* satisfies *D*_*A*(*i*)_ = 1. We refer to such a design as *disease-selected*.

Definition 4.2.1 appears innocuous but has a decisive consequence developed in Proposition 4.3.18: within study *i*, the variable *D*_*A*(*i*)_ is degenerate, and therefore carries zero information about association.

#### 4.2.2 Curation

Three classes of irregularity were resolved prior to analysis.

1. **Compound cells**. Entries such as 29(with depression) and 6.1/21 score mix a numeric value with a qualifier. The leading numeral was extracted where a unique numeric reading exists and the qualifier retained in a parallel annotation field; entries admitting no unique numeric reading were coded missing.
2. **Missingness coding**. The corpus uses NR, blank, and NaN interchangeably. These were unified, with the important semantic caveat that NR (“not reported”) denotes a quantity the study did not ascertain, not a quantity ascertained to be zero. This distinction is revisited in Section 7.
3. **Non-canonical cohort labels**. Rows labelled, e.g., “POTS in kids” or “Nausea/Vomiting” were flagged as subgroups and excluded from the primary analysis, entering only a sensitivity variant.

##### Definition 4.2.2

(Count reconstruction). Given a reported percentage *p*_*i*_ and sample size *n*_*i*_, the event count used in (5) is

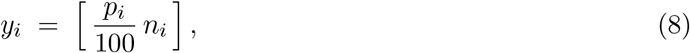

where [·] denotes rounding to the nearest integer, truncated to {0, …, *n*_*i*_}.

Equation (8) inverts a lossy transformation. A percentage reported to one decimal place determines *y*_*i*_ exactly whenever *n*_*i*_ *<* 1000, but for the largest cohorts (*n*_*i*_ ∼ 2*×*10^4^) the reported precision admits a range of integer counts. The induced error is *O*(*n*_*i*_ *×* 10^−3^) in the count and *O*(10^−3^) in the proportion, negligible relative to *τ* . Section 5.6 conducts an explicit rounding-sensitivity analysis where the conclusion depends on exact integer recovery.

#### 4.2.3 Data availability by estimand

The corpus is sparse. Overall symptom-cell completeness is 28%: of 22*×*29 possible study–symptom cells, fewer than one in three is populated. The median study reports 8 symptoms; the largest fully complete submatrix is 4 studies *×* 5 symptoms. Only one symptom (migraine/headache) is reported by as many as 15 studies, and only five symptoms by ten or more.

Table 1 summarizes the evidence available for each of the six directed comorbidity estimands. The imbalance is severe: *π*_POTS|hEDS_ is supported by five studies totalling *n* = 6343, whereas *π*_POTS|MCAS_ and *π*_hEDS|MCAS_ rest on two studies totalling *n* = 561.

**Table 1.** Evidence available per directed estimand. *k* denotes the number of contributing studies and *N* the aggregate sample size across those studies.

| Estimand | $k$ | $N$ |
| --- | --- | --- |
| $\pi_{\text{hEDS}\vert \text{POTS}}$ | 3 | 5473 |
| $\pi_{\text{MCAS}\vert \text{POTS}}$ | 2 | 5434 |
| $\pi_{\text{POTS}\vert \text{hEDS}}$ | 5 | 6343 |
| $\pi_{\text{MCAS}\vert \text{hEDS}}$ | 3 | 6202 |
| $\pi_{\text{POTS}\vert \text{MCAS}}$ | 2 | 561 |
| $\pi_{\text{hEDS}\vert \text{MCAS}}$ | 2 | 561 |
| $\mathbb{P}(\text{all three})$ | 1 | 8 |

The final row of Table 1 is consequential. A single study reports the triple co-occurrence. No meta-analytic pooling is possible for this quantity; it is reported as a single-study figure throughout, and never as a pooled estimate. Section 5.6 shows that this same study nonetheless yields the most informative single result in the corpus.

#### 4.2.4 Patient-level supplement

A secondary file by Weinstock et al. (2023) records 8 patients with 13 symptom indicators and three diagnostic indicators, coded ternary (Yes/unclear/possible). We treat unclear as unascertained rather than absent; the resulting ascertainment rate is 52% (54 of 104 patient–symptom cells). All eight patients carry an MCAS diagnosis, and all eight are recorded positive for anxiety/depression; these variables are therefore degenerate in this sample. Section 5.5 establishes that this supplement cannot support latent class analysis and quantifies what would be required.

The supplement additionally records generalized joint hypermobility, the one symptom that the biological account regards as approximately hEDS-specific rather than convergent across the triad. This variable is absent from the aggregate corpus and is therefore unavailable to Models A and B, whose symptom set consists entirely of convergent (multi-pathway) manifestations. Within the supplement, hypermobility is ascertained in five of eight patients and present in all five, but with near-universal triad membership at *N* = 8 it exhibits no discriminating variance; its specificity for hEDS can therefore be neither confirmed nor refuted here. We record this as a scope limitation: the analysis quantifies the evidential content of convergent symptoms precisely because the one putatively disease-specific symptom is unrepresented in the poolable data, and testing its specificity would require its systematic extraction in a future cohort.

### 4.3 Methodology

#### 4.3.1 Model A: hierarchical comorbidity meta-analysis

Pooling proportions across studies requires care: the variance of a proportion depends on its mean, and naive transformations are known to distort the pooled estimate (Barendregt et al., 2013; Schwarzer et al., 2019).

For each ordered pair (*A, B*), *A≠ B*, let *I*_*A*_ index the studies with *A*(*i*) = *A* reporting the prevalence of *B*. The model is (5)–(6) with priors (7). The joint posterior density is, up to a normalizing constant,

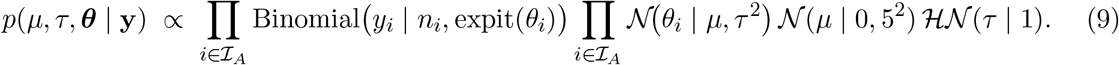

##### Proposition 4.3.1

(Reduction to a two-dimensional posterior). *The study-level effects θ*_1_, …, *θ*_*k*_ *can be integrated out exactly, leaving a posterior over* (*µ, τ*) *alone:*

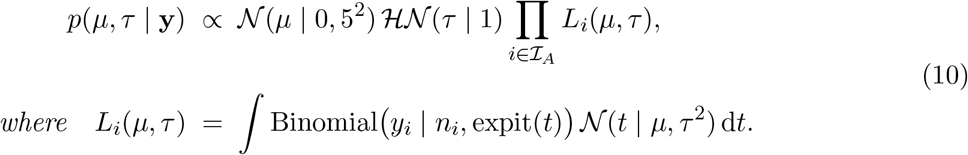

*Proof*. The *θ*_*i*_ are conditionally independent given (*µ, τ*) and each appears in exactly one likelihood factor, so the *k*-fold integral in *p*(*µ, τ* | **y**) ∝ *p*(*µ, τ*) *p*(**y** | ***θ***)*p*(***θ*** | *µ, τ*) d***θ*** factorizes into the product of *k* one-dimensional integrals *L*_*i*_.

Each *L*_*i*_ is a smooth one-dimensional integral against a Gaussian weight and is evaluated to machine precision by 40-node Gauss–Hermite quadrature. The right-hand side of (10) is then tabulated on a 161 *×* 121 grid over (*µ, τ*) ∈ [−6, 6] *×* (0, 4] and normalised by summation. Posterior summaries are read off the grid directly, and draws for downstream propagation are obtained by sampling grid cells in proportion to their mass with uniform jitter within each cell.

This replaces the Markov chain Monte Carlo scheme that a model of this form would ordinarily invoke. Because the parameter of scientific interest is only two-dimensional once Proposition 4.3.1 is applied, deterministic quadrature is not merely adequate but preferable: it is exactly reproducible, requires no burn-in, thinning, proposal tuning, or convergence diagnostics, and eliminates Monte Carlo error from the estimates entirely. We verified against a component-wise Metropolis-within-Gibbs implementation that the two agree to within 0.5 percentage points on every estimand—within the Monte Carlo error of the sampler itself—and report the deterministic results throughout.

Where *k* = 1 the hierarchical model is inestimable—no between-study variance exists to be identified—and we substitute the single-study Jeffreys posterior 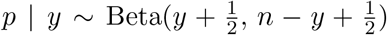, reporting the result as single-study rather than pooled.

#### 4.3.2 Model B: symptom-conditioned prediction

Model B addresses (Q2). By Lemma 4.1.5 the update requires the prior odds—supplied by Model A—and the likelihood ratios (2). The difficulty lies entirely in the denominator ℙ(*S* | *A, ¬B*), which is not directly reported by any study.

##### The reference-group problem

A common expedient is to approximate ℙ(*S* | *A, ¬B*) by the prevalence of *S* in cohorts selected on the *other* diagnoses. This is misspecified: the reference class for a patient known to have *A* is other patients with *A*, not patients with unrelated diagnoses. The resulting likelihood ratio answers a question nobody asked.

We instead exploit the fact that a disease-*A* cohort is itself a mixture of *B*-comorbid and non-comorbid patients, with mixing weight given precisely by the Model A estimand.

###### Lemma 4.3.2

(Mixture decomposition and deconvolution). *For any symptom S and distinct diagnoses A, B*,

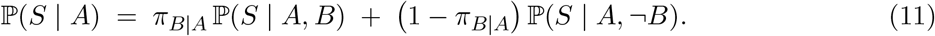

*Consequently, whenever π*_*B*|*A*_ *<* 1,

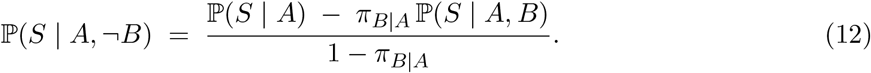

*Proof*. Equation (11) is the law of total probability applied to the partition *{B, ¬B}* of the sample space conditional on *A*, with weights ℙ(*B* | *A*) = *π*_*B*|*A*_ and ℙ(*¬B* | *A*) = 1 − *π*_*B*|*A*_. Solving for the second summand gives (12), the division being legitimate under the stated hypothesis.

The right-hand side of (12) involves ℙ(*S* | *A*), estimated by pooling symptom prevalences over *A*-cohorts; *π*_*B*|*A*_, estimated in Model A; and ℙ(*S* | *A, B*), which remains unobserved. We close the system with the following approximation.

###### Assumption 4.3.3

(Symptom expression dominated by the target diagnosis). ℙ(*S* | *A, B*) ≈ ℙ(*S* | *B*); that is, among patients carrying both diagnoses, the prevalence of *S* is approximated by its prevalence in *B*-selected cohorts.

###### Remark 4.3.4

(Status of Assumption 4.3.3). This is a genuine approximation, not an identity, and it is the weakest link in Model B. It would hold exactly if *S* **╨** *D*_*A*_ | *D*_*B*_. It is adopted because ℙ(*S* | *A, B*) is unreported in every study in the corpus; the alternative is to abandon (Q2) entirely.

Its plausibility is assessed indirectly in Section 5.2, where its failure manifests as infeasibility.

##### Feasibility and the coherence constraint

Equation (12) does not automatically return a probability. The constraint that governs when it does is entirely elementary—it is the observation that a joint probability cannot exceed its marginal—but it has consequences the literature has not drawn.

###### Proposition 4.3.5

(Feasibility constraint). *The quantity defined by* (12) *lies in* [0, 1] *if and only if*

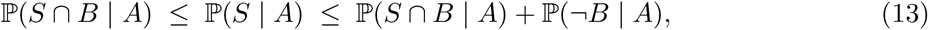

*where* ℙ(*S* ∩ *B* | *A*) = *π*_*B*|*A*_ℙ(*S* | *A, B*). *The left inequality is monotonicity under the inclusion S* ∩ *B* ⊆ *S; the right is the same statement applied to ¬B*.

*Proof*. Write *r* = ℙ(*S* | *A, ¬B*) as in (12), with numerator *ν* = ℙ(*S* | *A*) − *π*_*B*|*A*_ℙ(*S* | *A, B*) and positive denominator 1 − *π*_*B*|*A*_. Then *r* ≥ 0 iff *ν* ≥ 0 and *r* ≤ 1 iff *ν* ≤ 1 − *π*_*B*|*A*_, which are respectively the left and right inequalities of (13) after substituting *π*_*B*|*A*_ℙ(*S* | *A, B*) = ℙ(*S* ∩ *B* | *A*) by the multiplication rule.

###### Remark 4.3.6

(On the elementary character of (13)). Constraints of this form are the two-event case of the classical Fréchet–Hoeffding bounds, and could be presented in that language. We deliberately do not, because the substance of the result lies not in its depth—it is a one-line consequence of monotonicity—but in the empirical fact, established in Section 5.2, that separately published and separately pooled quantities routinely violate it. A violated elementary inequality is a stronger indictment of the evidence base than a violated sophisticated one.

###### Definition 4.3.7

(Incoherence fraction). Let 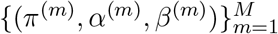 be posterior draws of (*π*_*B*|*A*_, ℙ(*S* | *A*), ℙ(*S* | *B*)) obtained from Model A and the symptom pooling. Define the deconvolved reference draw *r*^(*m*)^ = (*α*^(*m*)^ − *π*^(*m*)^*β*^(*m*)^)*/*(1 − *π*^(*m*)^). The *incoherence fraction* of the cell (*S*; *B* | *A*) is

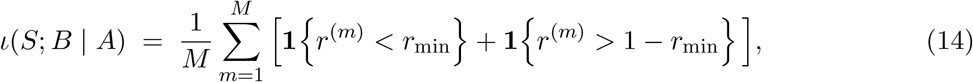

with floor *r*_min_ = 0.01. Both tails are counted, because Proposition 4.3.5 is a two-sided constraint: *r*^(*m*)^ *>* 1 is as infeasible as *r*^(*m*)^ *<* 0, and is the more common failure in this corpus.

###### Remark 4.3.8

(Interpretation). By Proposition 4.3.5, *r*^(*m*)^ ∈*/* [0, 1] certifies that the triple (*π*^(*m*)^, *α*^(*m*)^, *β*^(*m*)^) cannot arise from any joint distribution satisfying Assumption 4.3.3. Since each component is separately a legitimate posterior draw from a well-specified marginal model, the incompatibility must lie in their *conjunction*. Two explanations are available and are not mutually exclusive: (i) Assumption 4.3.3 fails, so ℙ(*S* | *A, B*)*≠* ℙ(*S* | *B*); or (ii) the three quantities, being pooled over disjoint study sets with heterogeneous ascertainment, are not estimates of parameters of a common population. Either way, *ι* is a valid diagnostic of internal inconsistency, and cells with large *ι* must be excluded rather than clipped.

Cells are retained only if *ι* ≤ 0.25 and both contributing symptom estimates rest on at least two studies. Surviving likelihood ratios are capped to [10^−1^, 10] to prevent a single sparsely reported symptom from dominating the product—a form of explicit regularization whose effect is documented rather than concealed.

##### Tempered updating under dependence

Remark 4.1.6 noted that (3) is implausible. Correlated symptoms constitute partially redundant evidence, so the product in (4) overstates the total information. We therefore report, alongside the naive update, a tempered variant

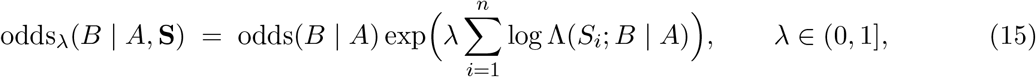

with *λ* = 1 (naive) and 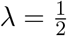 (tempered).

###### Remark 4.3.9

(Status of *λ*). Equation (15) is deliberately crude: it simply halves the total log-evidence when 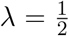, on the reasoning that *n* correlated symptoms convey less than *n* symptoms’ worth of independent information. We resist dressing this as an estimator. The device is a *sensitivity analysis*, not a correction: *λ* is not estimated and cannot be, since identifying it would require exactly the joint symptom distribution whose absence motivates it. Its only role is to bracket the answer. The reader should treat the gap between the *λ* = 1 and 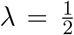 results as a lower bound on the uncertainty attributable to symptom dependence, and neither endpoint as a calibrated estimate. Formal machinery for down-weighting misspecified likelihoods exists and would supply a principled *λ* given data we do not have; invoking it here would lend the choice an authority it has not earned.

##### Uncertainty propagation

All quantities entering (15) are represented by posterior draws. For each Monte Carlo replicate *m* we draw *π*^(*m*)^ from the Model A posterior and *α*^(*m*)^, *β*^(*m*)^ from the corresponding symptom posteriors, compute *r*^(*m*)^ by (12), form Λ^(*m*)^, and evaluate (15). The reported estimate is the posterior median of {ℙ^(*m*)^(*B* | *A*, **S**)} and the reported interval its 2.5^th^ and 97.5th percentiles, with *M* = 4000. No delta-method or asymptotic approximation is used; the propagated distribution is exact up to Monte Carlo error.

##### Construction of the evaluation panel

The two-sided coherence screen of Definition 4.3.7 retains between four and ten symptoms per direction, out of the twenty-nine recorded in the corpus and the fourteen to nineteen that are reported for both members of a given pair. Exhaustive evaluation over all retained symptoms would require 2^10^ = 1024 subsets per direction, which is computationally feasible but yields accumulation curves too dense to interpret and confers a false impression of resolution given the width of the underlying intervals. We therefore fix a *panel* of the *κ* = 4 retained symptoms with the largest posterior median likelihood ratio, and enumerate its 2^4^ = 16 subsets. The panel size is set by the scarcest direction: after the two-sided coherence screen of Definition 4.3.7, the direction ℙ(MCAS | hEDS) retains only four symptoms, and a common panel size is required if the six directions are to be compared on equal terms.

###### Remark 4.3.10

(Status of the panel size). The choice *κ* = 4 is forced by the scarcest direction, not chosen for tractability. Between zero and six retained symptoms per direction are therefore excluded from the panel despite passing the coherence screen. Because the excluded symptoms are by construction those with likelihood ratios closest to unity, their omission is conservative: including them would move the posteriors further from the prior under naive updating, and the direction of that movement is precisely the quantity the conditional-independence assumption cannot be trusted to estimate. Every figure reported below as “all four symptoms” therefore means *all four panel symptoms*, not all symptoms recorded for that direction.

##### Pathway-structured evidence

The symptom list is not an unstructured collection: each symptom is attributable, on pathophysiological grounds, to one of six biological pathways—connective-tissue, autonomic, mast-cell/inflammatory, gastrointestinal, neurovascular, and central-sensitisation. Denote by *P* = *{P*_1_, …, *P*_6_} the induced partition of the symptom set *S*, so that each *S*_*j*_ belongs to exactly one *P*_*ℓ*_. The biological premise—that a symptom drawn from a pathway distinct from the primary diagnosis’s characteristic system constitutes evidence of a *second* underlying process— motivates the following contrast.

###### Definition 4.3.11

(Within- and cross-pathway panels). For a target direction (*A, B*) and an integer *κ* ≥ 2, a *within-pathway panel* is a set **S**_w_ ⊆ *P*_*ℓ*_ of *κ* symptoms drawn from a single pathway *P*_*ℓ*_; a *cross-pathway panel* **S**_c_ is a set of *κ* symptoms drawn from *κ* distinct pathways. Both are evaluated through the tempered update (15) with 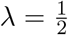, holding *κ* fixed so that the comparison isolates the effect of pathway *dispersion* from that of panel *size*.

The quantity of interest is the pathway-dispersion contrast Δ_*B*|*A*_ = ℙ(*B* | *A*, **S**_c_) − ℙ(*B* | *A*, **S**_w_), positive values of which indicate that multi-system symptom evidence is more diagnostic of comorbid disease than an equal quantity of single-system evidence, as the biological account predicts.

##### Decision-analytic layer

Questions (Q1)–(Q3) concern estimation; the clinical motivation (Q4) concerns a *decision*. We adopt a threshold rule, and we are explicit that this is not a decision analysis: no loss function is specified, no cost of surveillance or of missed diagnosis is available, and the thresholds below are asserted rather than derived. What follows should be read as a reporting device that renders the posteriors comparable across directions, not as an optimal policy.

###### Definition 4.3.12

(Monitoring rule). Fix a threshold *t* ∈ (0, 1). Under symptom profile **S**, direction (*A, B*) triggers a *monitoring recommendation* if the posterior median satisfies 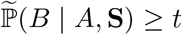. For a panel of size *n*, define the *threshold-crossing number* 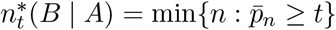, where 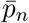 is the posterior median averaged over all *κ* = *n* symptom subsets of the direction’s panel, and 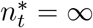 if the panel never reaches *t*.

We report 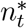 for *t* ∈ {0.30, 0.50}, values chosen to bracket plausible low- and moderate-suspicion monitoring thresholds; the analysis is threshold-agnostic in that the full curve 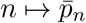 is displayed, allowing any reader to substitute an alternative *t*. We stress that *t* encodes an implicit cost ratio between missed comorbidity and unnecessary investigation that we do not attempt to estimate; the layer therefore quantifies *how much symptom evidence is required* to reach a given level of suspicion, not *whether* that level is the correct one.

#### 4.3.3 Model C: latent structure

##### Definition 4.3.13

(Latent class model). Let *S*_1_, …, *S*_*J*_ be binary indicators. The *K*-class latent class model posits a latent *C* ∈ {1, …, *K}* with ℙ(*C* = *c*) = *π*_*c*_ such that the indicators are conditionally independent given *C*:

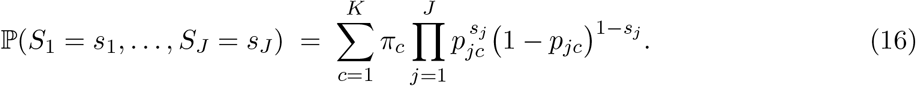

The parameter vector is *ϕ* = (***π*, P**) with (*K* − 1) + *KJ* free components.

Estimation proceeds by expectation–maximization (Dempster et al., 1977). Writing 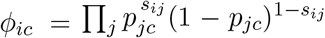 for the class-*c* likelihood of patient *i*’s response pattern, the E-step responsibilities are

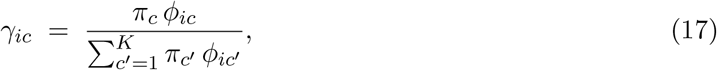

computed in log-space with a max-subtraction stabilization; the M-step updates are

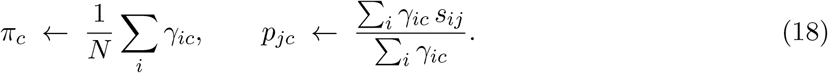

Model order is selected by 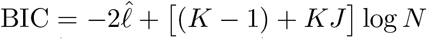, and classification sharpness by the normalised entropy 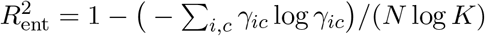.

The critical question is not whether (17)–(18) can be executed—they always can—but whether *ϕ* is identified by the available observables.

##### Theorem 4.3.14

(Non-identifiability of the latent class model from univariate marginals). *Let J* ≥ 2 *and suppose*) *the observation functional is the vector of univariate marginals T* (*P*) = ℙ(*S*_1_ = 1), …, ℙ(*S*_*J*_ = 1) . *Then for every K* ≥ 2 *the parameter ϕ is not identified by T* ; *and the pair* (*K, ϕ*) *is not identified by T even when K* = 1 *is admitted. The restriction to K* ≥ 2 *is necessary: at K* = 1 *the model is the product measure, T is the identity on ϕ* = (*p*_1_, …, *p*_*J*_), *and ϕ* is *identified. Specifically, there exist joint distributions P*_1_*≠ P*_2_ *on {*0, 1}^*J*^ *with T* (*P*_1_) = *T* (*P*_2_) *but with different conditional dependence structure*.

*Proof*. It suffices to exhibit the counterexample for *J* = 3; the construction extends to larger *J* by taking products with arbitrary fixed marginals. Let *P*_1_ be the product measure with 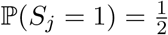 independently, so 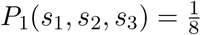 for all (*s*_1_, *s*_2_, *s*_3_) ∈ {0, 1}^3^. Let *P*_2_ place mass 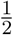 on (0, 0, 0) and mass 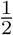 on (1, 1, 1), and zero elsewhere. Then for each *j*,

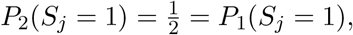

so *T* (*P*_1_) = *T* (*P*_2_). However under *P*_1_ the indicators are mutually independent, whereas under *P*_2_ they are almost surely equal, so *P*_1_ ≠ *P*_2_. Since *P*_1_ is realized by (16) with *K* = 1 and *p*_*j*1_ = ^1^, while *P*_2_ is realized with 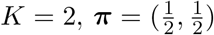, *p*_*j*1_ = 0, *p*_*j*2_ = 1, two distinct parameter values induce identical observables, establishing non-identifiability of (*K, ϕ*). For fixed *K* ≥ 2 the same conclusion follows more directly: setting *p*_*jc*_ = *p*_*j*_ for all *c* makes the likelihood independent of ***π***, so every ***π*** on the simplex yields identical observables.

##### Corollary 4.3.15

(Consequence for the present corpus). *The aggregate corpus reports, for each cohort, only univariate symptom marginals. By Theorem 4.3.14 no latent class model with K* ≥ 2*— that is, no model positing latent heterogeneity at all—is identified from these data. Any numerical output of* (17)*–*(18) *applied to such data is determined by the initialization and the stopping rule rather than by the evidence*.

##### Remark 4.3.16

(On the elementary character of Theorem 4.3.14). The theorem is a restatement, in latent-class language, of the elementary fact that univariate marginals do not determine a joint law—equivalently, that they leave the copula free. We claim no depth for it. It is stated formally because the corollary it supports is routinely violated in practice: latent class models are fitted to exactly these data and their output reported as structure.

##### Remark 4.3.17

(What would suffice). Theorem 4.3.14 concerns univariate marginals. Classical identifiability results for finite mixtures of Bernoulli products (Allman et al., 2009; Kruskal, 1977) establish that trivariate marginals of *J* ≥ 3 conditionally independent indicators generically suffice to identify a two-class model. The obstruction here is thus specifically the absence of joint—not merely the paucity of marginal—information.

We next show that the obstruction is not remediable by collecting more studies of the same design.

##### Proposition 4.3.18

(Degeneracy under disease-selected sampling). *Let study i be disease-selected on A*(*i*) = *A in the sense of Definition 4*.*2*.*1*. *Then within study i the indicator D*_*A*_ *is almost surely constant*, Var(*D*_*A*_) = 0, *and the trivariate table of* (*D*_POTS_, *D*_hEDS_, *D*_MCAS_) *is not observable from study i. Consequently, no collection of disease-selected studies, however large, identifies the joint law of* (*D*_POTS_, *D*_hEDS_, *D*_MCAS_) *in an unselected population*.

*Proof*. By construction *D*_*A*_ = 1 almost surely on the sampling frame of study *i*, so its variance is zero and its marginal carries no information. The observable content of study *i* concerning the remaining two indicators is the conditional law given *D*_*A*_ = 1, i.e. a slice of the trivariate table, not the table itself. A union of such slices over *A* ∈ *D* supplies the three conditional laws *{L*(*D*_*B*_, *D*_*C*_ | *D*_*A*_ = 1)} but never the cell ℙ(*D*_POTS_ = 0, *D*_hEDS_ = 0, *D*_MCAS_ = 0), which lies outside every sampling frame. Since the unselected joint law assigns positive mass to that cell in general, it is not determined by the observables.

Proposition 4.3.18 is a selection-bias result of a particularly clean kind: the missing cell is missing by design, not by chance, and its absence is not detectable from within the data. It implies that the latent-structure question (Q3) cannot be settled by any meta-analysis of disease-selected cohorts, and requires either an unselected population sample or an explicit, externally justified model of the selection mechanism.

#### 4.3.4 Recovery of a joint contingency table

Although Proposition 4.3.18 precludes recovery of the trivariate table, a weaker but still valuable object is available when a single cohort reports all three diagnoses *and* their triple intersection.

##### Proposition 4.3.19

(Exact recovery by inclusion–exclusion). *Let a cohort of size N be selected on diagnosis A, so that D*_*A*_ = 1 *throughout, and suppose it reports the marginal proportions* 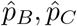 *of the other two diagnoses together with the proportion* 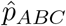 *carrying all three. Then, writing* 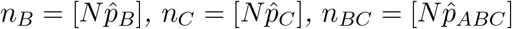, *the full* 2 *×* 2 *contingency table of D*_*B*_ *× D*_*C*_ *within the cohort is determined:*

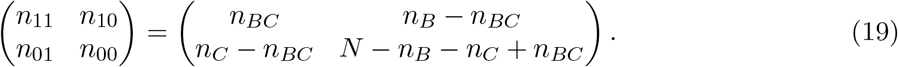

*Proof*. Since *D*_*A*_ ≡ 1 on the cohort, the event “all three” coincides with *{D*_*B*_ = 1, *D*_*C*_ = 1}, giving *n*_11_ = *n*_*BC*_. The remaining cells follow from the marginal constraints *n*_11_ + *n*_10_ = *n*_*B*_ and *n*_11_ + *n*_01_ = *n*_*C*_, and from ∑*n*_*ab*_ = *N*, which yields *n*_00_ = *N* −*n*_10_ −*n*_01_ −*n*_11_ = *N* −*n*_*B*_ −*n*_*C*_ + *n*_*BC*_, the inclusion–exclusion identity.

Given (19), the hypothesis *D*_*B*_ **╨** *D*_*C*_ within the cohort is testable by Fisher’s exact test (1922), with the conditional odds ratio 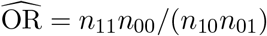 and Woolf standard error (1955) 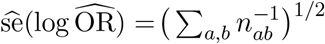.

## 5 Results

### 5.1 Model A: pooled directed comorbidity

Table 2 reports the six directed comorbidity estimands. Each entry is the posterior median of *π*_*B*|*A*_ = expit(*µ*) with its 95% credible interval, the posterior median of the heterogeneity standard deviation *τ*, the 95% prediction interval obtained by drawing *θ*_new_ ∼ *N* (*µ, τ* ^2^) and transforming, the number of contributing studies *k*, and the pooled sample size.

**Table 2.** Model A: Bayesian hierarchical estimates of the six directed comorbidity probabilities. Credible intervals (CrI) are 95% posterior intervals for the pooled proportion; prediction intervals (PrI) are 95% intervals for the true proportion in a new study. *τ* is the posterior median between-study standard deviation on the logit scale.

| Estimand | Median (%) | 95% CrI | 95% PrI | $\tau$ | $k$ | $N$ |
| --- | --- | --- | --- | --- | --- | --- |
| $\pi_{\text{hEDS}\vert \text{POTS}}$ | 12.1 | [3.5, 38.5] | [0.9, 70.8] | 1.02 | 3 | 5473 |
| $\pi_{\text{MCAS}\vert \text{POTS}}$ | 3.9 | [0.7, 21.6] | [0.2, 55.1] | 1.15 | 2 | 5434 |
| $\pi_{\text{POTS}\vert \text{hEDS}}$ | 46.6 | [32.5, 61.5] | [17.8, 78.9] | 0.63 | 5 | 6343 |
| $\pi_{\text{MCAS}\vert \text{hEDS}}$ | 32.2 | [17.6, 55.5] | [7.8, 75.5] | 0.55 | 3 | 6202 |
| $\pi_{\text{POTS}\vert \text{MCAS}}$ | 49.5 | [22.0, 77.2] | [11.4, 88.7] | 0.44 | 2 | 561 |
| $\pi_{\text{hEDS}\vert \text{MCAS}}$ | 27.6 | [10.3, 69.4] | [4.2, 86.4] | 0.65 | 2 | 561 |

Three features warrant comment. First, the estimates are markedly *asymmetric*: *π*_POTS|hEDS_ = 46.6% exceeds *π*_hEDS|POTS_ = 12.1% by nearly a factor of four. By (1) this is the expected consequence of differing marginal prevalences and carries no directional-causal content; it would be an error, common in the clinical literature, to read the larger number as evidence that hEDS “causes” POTS more than the reverse. Second, the credible intervals are wide precisely where *k* is small. The two directions resting on *k* = 2 studies (*π*_POTS|MCAS_, *π*_hEDS|MCAS_) have the widest intervals; the *π*_POTS|MCAS_ interval spans [11.9, 73.3], an admission of near-ignorance. This is the intended behaviour of the weakly-informative *τ* prior discussed in Section 4.3: with few studies the frequentist DerSimonian–Laird estimator would collapse 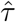 toward zero and produce a spuriously narrow interval, whereas the Bayesian treatment propagates heterogeneity uncertainty honestly. Third, the prediction intervals are substantially wider than the credible intervals in every row, and for the small-*k* directions they approach the entire unit interval. The prediction interval, not the credible interval, is the statistically appropriate summary of what a subsequent study might find, and its width here is a quantitative statement of how little the corpus constrains the phenomenon.

Figure 1 renders the six estimands as a directed graph, with edge weight encoding *k* and node size encoding pooled sample size. The visual dominance of the hEDS node reflects its provenance from a small number of large cohorts; the thin POTS *↔* MCAS edges mark the directions where evidence is weakest.

**Figure 1.**
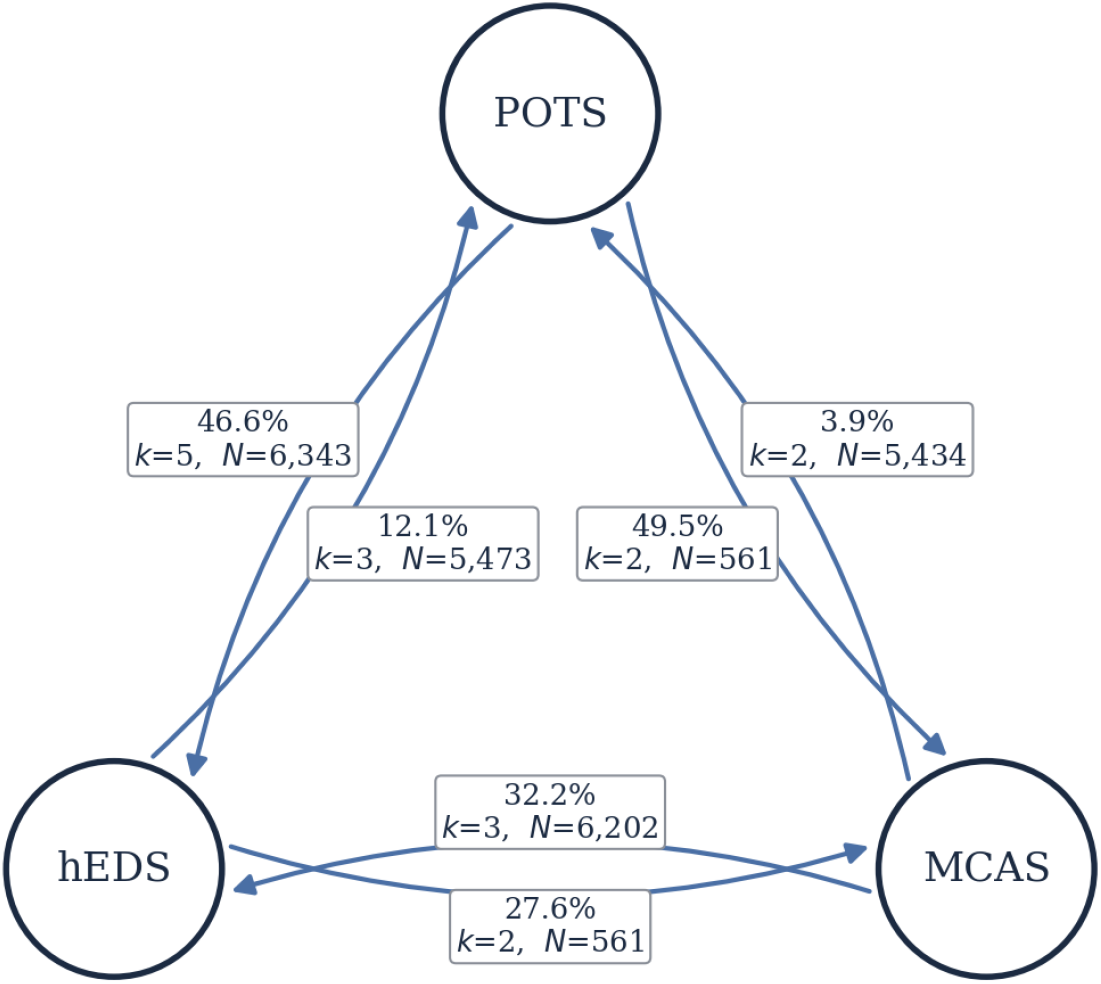
Directed comorbidity network. Each of the six directed estimands is drawn as its own arc, with its estimate printed on that arc, so that every figure is unambiguously attached to the arrow it describes. An arrow *A* → *B* denotes the pooled estimate of *π*_*B*|*A*_; *k* is the number of studies pooled for that direction; edge weight is proportional to *k* and node area to total study population. The asymmetry between the POTS–hEDS arcs and the thinness of the MCAS arcs are the graphical counterparts of Table 2.

The estimates in Table 2 inherit the reliability of their inputs, which is limited. All contributing cohorts are specialty-or referral-selected, so *π*_*B*|*A*_ estimates a comorbidity probability *within the referred population*, not in any general population. The direction of referral bias is not identifiable from the data; we return to this in Section 6.

### 5.2 Model B: symptom-conditioned prediction and the coherence diagnostic

#### 5.2.1 Coherence screening

Applying Definition 4.3.7 to all 102 evaluable symptom-by-direction cells, 60 cells (59%) are excluded from Model B. It is essential to separate the two grounds of exclusion, which are frequently conflated and which carry very different evidential weight. 26 cells (25.5%) are excluded for *incoherence* proper, i.e. *ι >* 0.25; a further 46 (45%) fail the separate requirement that both contributing symptom prevalences rest on at least two studies, and the two grounds overlap. The latter is a data-sufficiency rule, not a finding about internal consistency.

The decisive observation is that infeasibility is overwhelmingly *upper-tail*. Of the 26 incoherent cells, 21 are driven by posterior mass at *r >* 1 rather than *r <* 0. A one-sided diagnostic that tested only the lower Fréchet bound would report five incoherent cells and pass the remaining twenty-one into the evidence synthesis, where they would contribute likelihood ratios computed from a reference prevalence exceeding unity. We flag this because the one-sided screen is the natural first implementation, and it understates the problem by a factor of six.

A concrete instance of each tail makes the mechanism transparent. For the cell (hives; MCAS | hEDS) the pooled inputs are *π*_MCAS|hEDS_ = 0.33, ℙ(hives | MCAS) = 0.52, and ℙ(hives | hEDS) = 0.15. The lower Fréchet bound of (13) requires

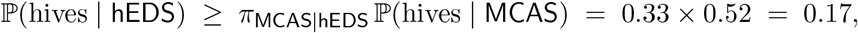

yet the reported left-hand side is smaller. No joint distribution exists in which at least 17% of hEDS patients have hives *via their MCAS comorbidity alone* while only 15% have hives in total. For the opposite tail, the cell (allergy symptoms; POTS | hEDS) has *ι* = 0.94 concentrated entirely at *r >* 1: the deconvolved prevalence of allergy symptoms among hEDS patients *without* POTS exceeds one, which is impossible.

Two explanations remain available and are not mutually exclusive, and the diagnostic cannot separate them. Either Assumption 4.3.3 fails, so that ℙ(*S* | *A, B*)*≠* ℙ(*S* | *B*); or the three pooled quantities, drawn from disjoint study sets with heterogeneous ascertainment, are not estimates of parameters of a common population. We regard the first as the more parsimonious reading in most instances, because *B*-selected cohorts are selected on *B*-characteristic symptoms and therefore report those symptoms at rates inflated relative to *B*-comorbid patients drawn from an *A*-cohort. On that reading the diagnostic is primarily an audit of Assumption 4.3.3 rather than an indictment of the literature, and we have withdrawn the stronger claim made in an earlier version of this work. Figure 2 displays the decomposition across cells.

**Figure 2.**
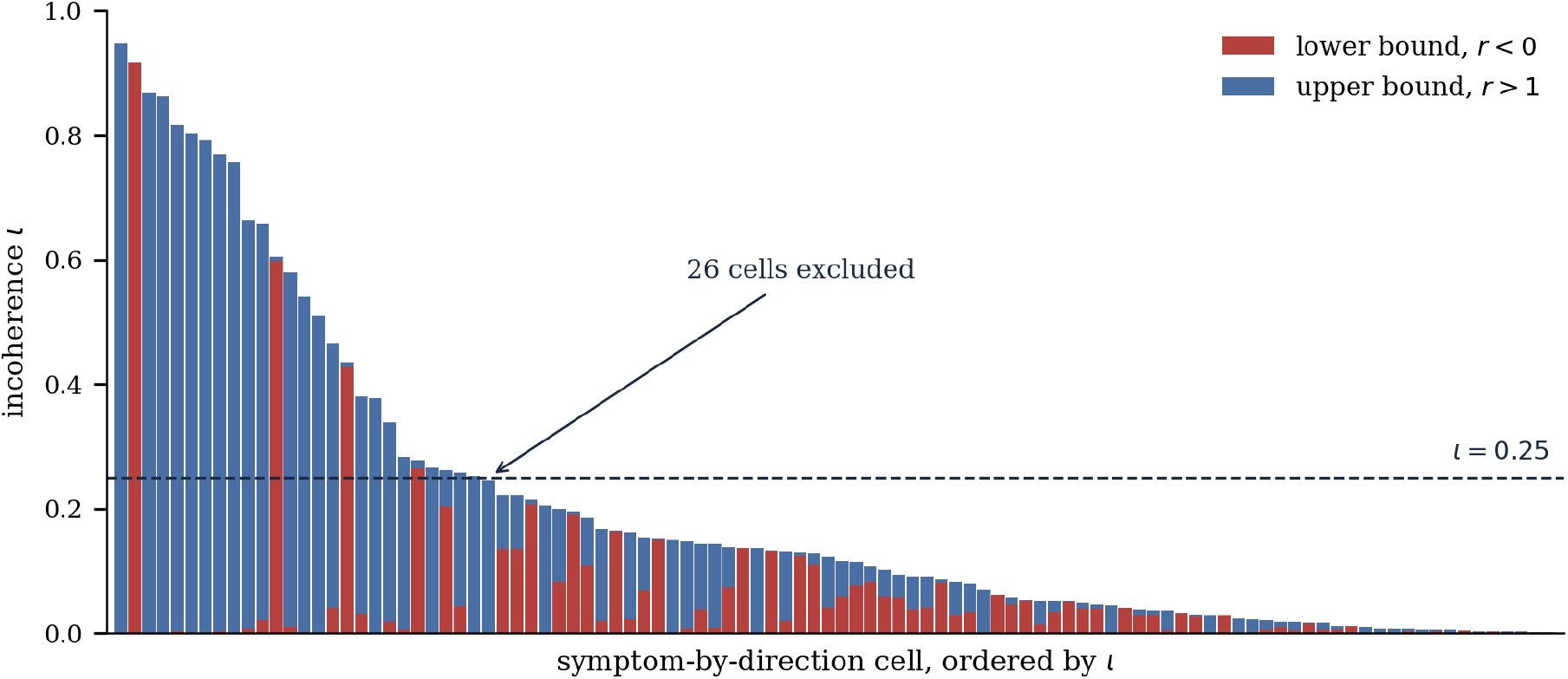
Two-sided coherence diagnostic. Each bar is the incoherence fraction *ι*(*S*; *B* | *A*) of (14), decomposed into posterior mass below the lower Fréchet bound (*r <* 0) and above the upper bound (*r >* 1); bars above the dashed threshold (*ι* = 0.25) mark cells excluded from Model B. Upper-tail infeasibility dominates, and would be invisible to a one-sided screen. Incoherence is a property of the *conjunction* of separately-valid pooled marginals.

##### Remark 5.2.1

(Epistemic significance, and its limits). The coherence diagnostic is, to our knowledge, an underused by-product of meta-analytic pooling. It requires no additional data: the mere act of combining marginals from disjoint study sets and checking the elementary inequalities (13) exposes internal contradictions that no single study could reveal. Outright infeasibility afflicts a quarter of the testable cells, and the diagnostic isolates them decisively while quantifying partial infeasibility elsewhere. We are, however, careful about what this licenses. The exclusions are a claim about specific symptom-by-direction quantities, not a global verdict on the literature, and they cannot distinguish failure of Assumption 4.3.3 from genuine incompatibility between study sets; on the more parsimonious reading the screen is an audit of that assumption.

#### 5.2.2 Likelihood ratios and evidence accumulation

For the retained cells, Figure 3 presents the posterior likelihood ratios Λ(*S*; *B* | *A*). Ratios are capped at 10 by the regularization of Section 4.3.2; credible intervals are wide, reflecting the small number of studies underlying each symptom prevalence.

**Figure 3.**
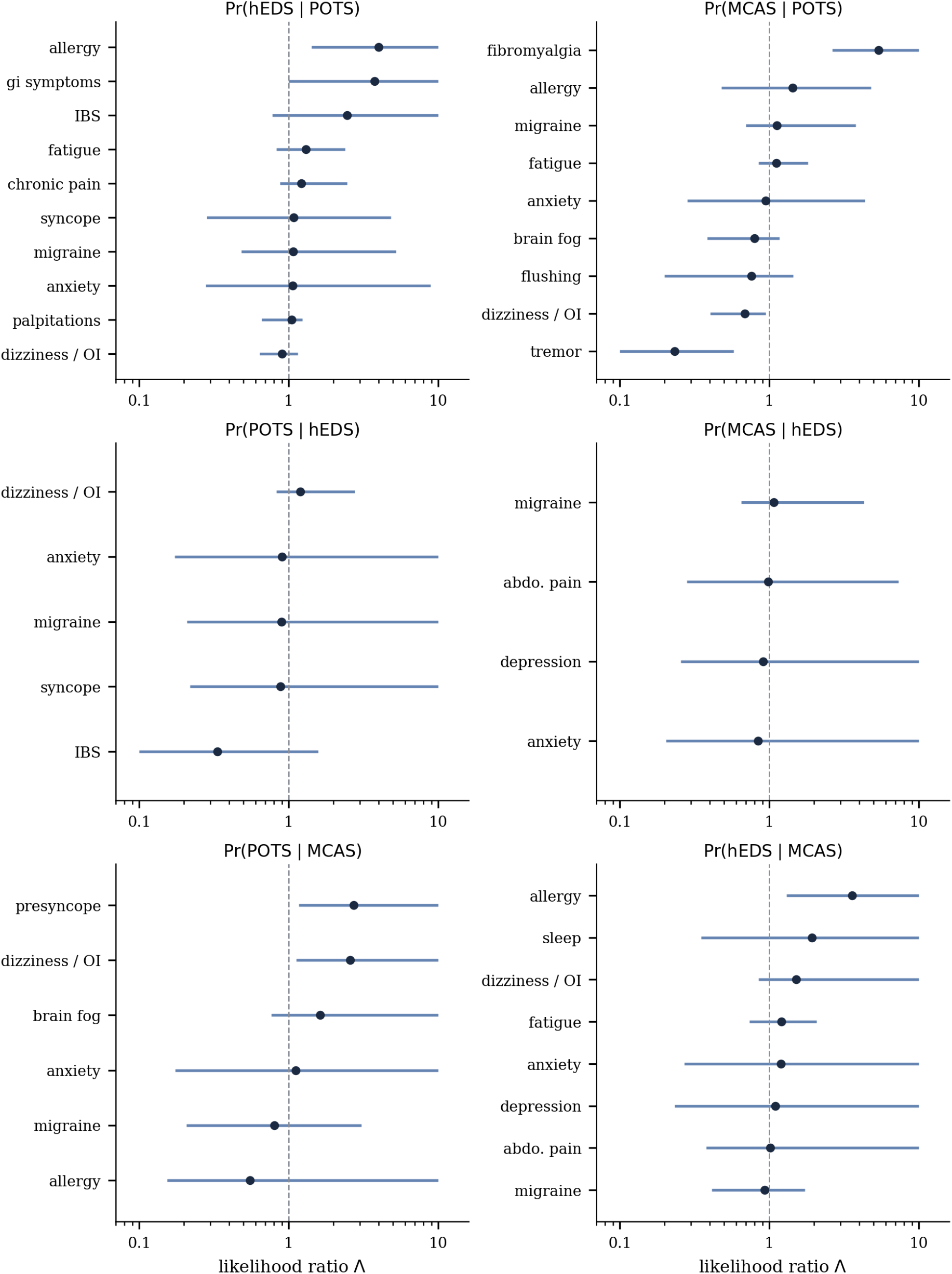
Symptom likelihood ratios by direction. Points are posterior medians of Λ(*S*; *B*|*A*); bars are 95% credible intervals; the vertical reference line at Λ = 1 separates evidence for (*>* 1) from evidence against (*<* 1) the target diagnosis. The logarithmic abscissa and the width of the intervals both underscore the limited resolving power of individual symptoms.

Table 3 summarizes the endpoints of evidence accumulation: the prior *π*_*B*|*A*_ (no symptoms), the posterior when all four panel symptoms are present under the naive update (*λ* = 1), and under the tempered update 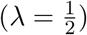. Figure 4 traces the full trajectory as symptoms accumulate.

**Table 3.** Model B: posterior probability of the target diagnosis given all four panel symptoms (Section 4.3.2), under naive (*λ* = 1) and tempered 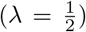 updating, compared with the symptom-free prior. All values are posterior medians (%). The symptom-free prior is the Model A estimand of Table 2 and agrees with it exactly; all downstream analyses draw from a single shared posterior sample (*M* = 2 *×* 10^4^) so that repeated quantities are numerically identical across tables.

| Direction | Prior | All six, $\lambda = 1$ | All six, $\lambda = \frac{1}{2}$ | 95% CrI ($\lambda = \frac{1}{2}$) |
| --- | --- | --- | --- | --- |
| $\mathbb{P}(\text{hEDS} \vert \text{POTS}, \mathbf{S})$ | 12.1 | 87.9 | 49.7 | [22, 82] |
| $\mathbb{P}(\text{MCAS} \vert \text{POTS}, \mathbf{S})$ | 3.9 | 30.0 | 11.9 | [4, 34] |
| $\mathbb{P}(\text{POTS} \vert \text{hEDS}, \mathbf{S})$ | 46.6 | 52.5 | 50.1 | [26, 80] |
| $\mathbb{P}(\text{MCAS} \vert \text{hEDS}, \mathbf{S})$ | 32.2 | 34.2 | 33.4 | [16, 65] |
| $\mathbb{P}(\text{POTS} \vert \text{MCAS}, \mathbf{S})$ | 49.5 | 95.6 | 82.1 | [53, 97] |
| $\mathbb{P}(\text{hEDS} \vert \text{MCAS}, \mathbf{S})$ | 27.6 | 85.0 | 59.8 | [32, 91] |
| bottomrule |  |  |  |  |

**Figure 4.**
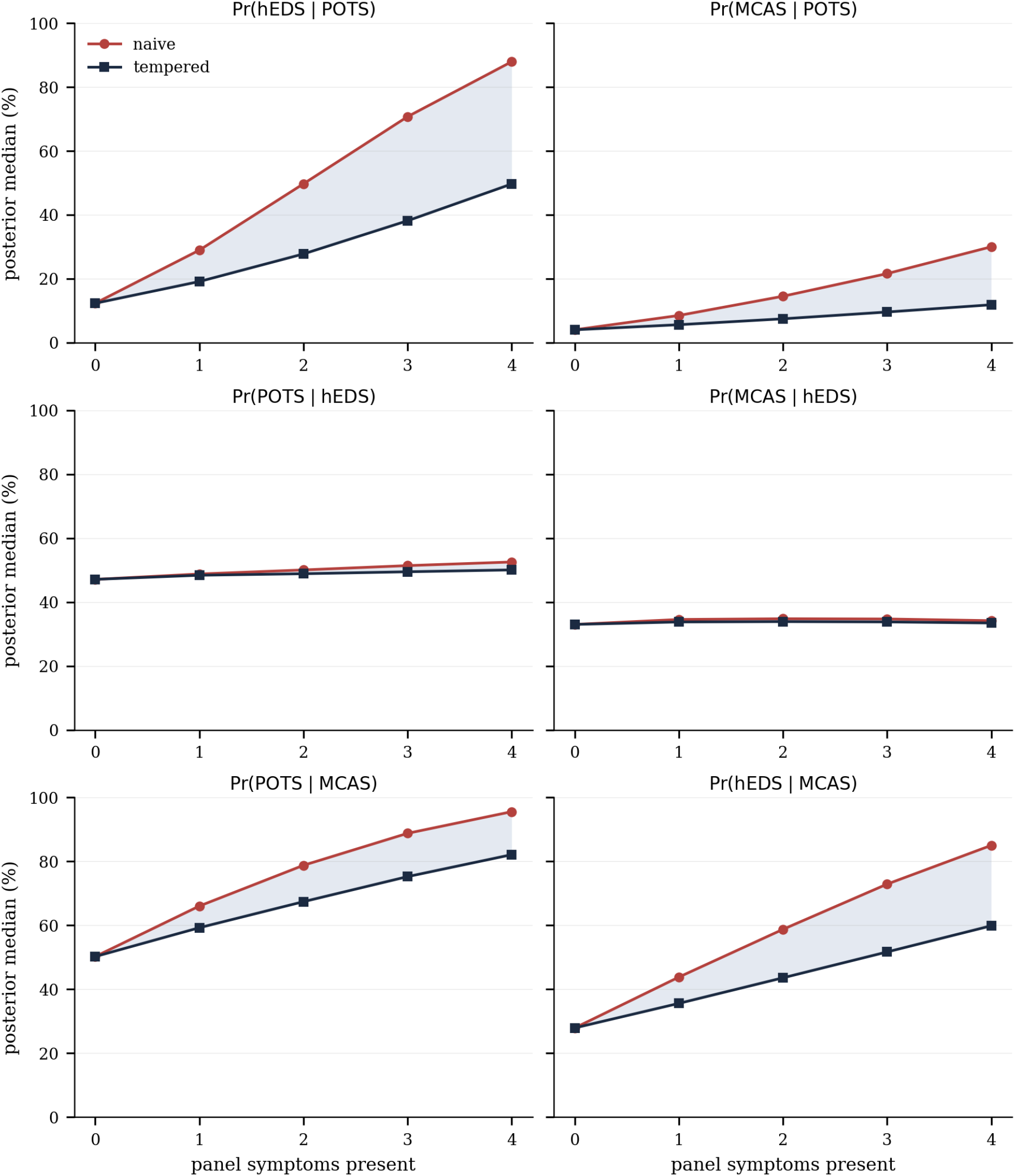
Posterior probability as symptom evidence accumulates, per direction. The upper (naive, *λ* = 1) and lower (tempered, 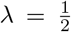) curves bound the effect of the conditional-independence assumption; their divergence widens with the number of symptoms, as (15) predicts. The shaded region between them is a lower bound on the analyst’s dependence-induced uncertainty, not a corrected estimate.

The gap between the two curves is the central cautionary finding of Model B. Under naive updating, sour symptoms drive ℙ(hEDS | POTS, **S**) from a prior of 12.1% to 87.9%; under tempered updating the same evidence yields 49.7%. Since no data in the corpus identifies *λ* (Remark following (15)), the honest report is the interval [49.7, 87.9], whose width is itself a measure of ignorance about symptom dependence. We therefore regard the tempered column of Table 3 as the defensible headline and the naive column as an upper bound obtained under an assumption known to be false.

### 5.3 Pathway structure: is multi-system evidence more diagnostic?

Table 4 and Figure 5 report the pathway-dispersion contrast Δ_*B*|*A*_ of Definition 4.3.11, with within- and cross-pathway panels matched at *κ* = 2 symptoms and evaluated at 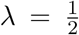. As with the evaluation panel, *κ* is forced downward by the two-sided screen: three directions retain no pathway containing three surviving symptoms, so *κ* = 3 is infeasible.

**Table 4.** Pathway-dispersion contrast. Posterior medians (%) for within-pathway and cross-pathway symptom panels of equal size (*κ* = 2), with the contrast Δ_*B*|*A*_ in percentage points. Positive Δ indicates multi-system evidence is more diagnostic than single-system evidence of equal size. Δ is computed as the median of the *paired* difference over shared posterior draws and therefore need not equal the difference of the two column medians, the median being non-linear. An asterisk marks the single contrast whose 95% credible interval excludes zero; it does so by 0.1 percentage points, and across six contrasts the expected number of 95% intervals excluding zero under the null is 0.3, so this should not be read as evidence of a pathway effect. No multiplicity adjustment is applied, and none would leave the contrast distinguishable from zero. Two directions admit only a single within-pathway pair, so their “within” column rests on one panel rather than an average.

| Direction | Prior | Within-pathway | Cross-pathway | $\Delta_{B\vert A}$ (pp) | 95% CrI on $\Delta$ |
| --- | --- | --- | --- | --- | --- |
| $\mathbb{P}(\text{hEDS} \mid \text{POTS}, \cdot)$ | 12.1 | 16.1 | 19.3 | +2.9 | [+0.2, +10.0]* |
| $\mathbb{P}(\text{MCAS} \mid \text{POTS}, \cdot)$ | 3.9 | 5.4 | 4.4 | −0.9 | [−5.3, −0.1]* |
| $\mathbb{P}(\text{POTS} \mid \text{hEDS}, \cdot)$ | 46.6 | 49.0 | 43.1 | −7.2 | [−30.8, +14.5] |
| $\mathbb{P}(\text{MCAS} \mid \text{hEDS}, \cdot)$ | 32.2 | 31.8 | 34.2 | +1.5 | [−20.1, +18.2] |
| $\mathbb{P}(\text{POTS} \mid \text{MCAS}, \cdot)$ | 49.5 | 64.4 | 58.2 | −5.9 | [−17.3, +7.0] |
| $\mathbb{P}(\text{hEDS} \mid \text{MCAS}, \cdot)$ | 27.6 | 36.0 | 37.7 | +1.7 | [−10.9, +14.9] |

**Figure 5.**
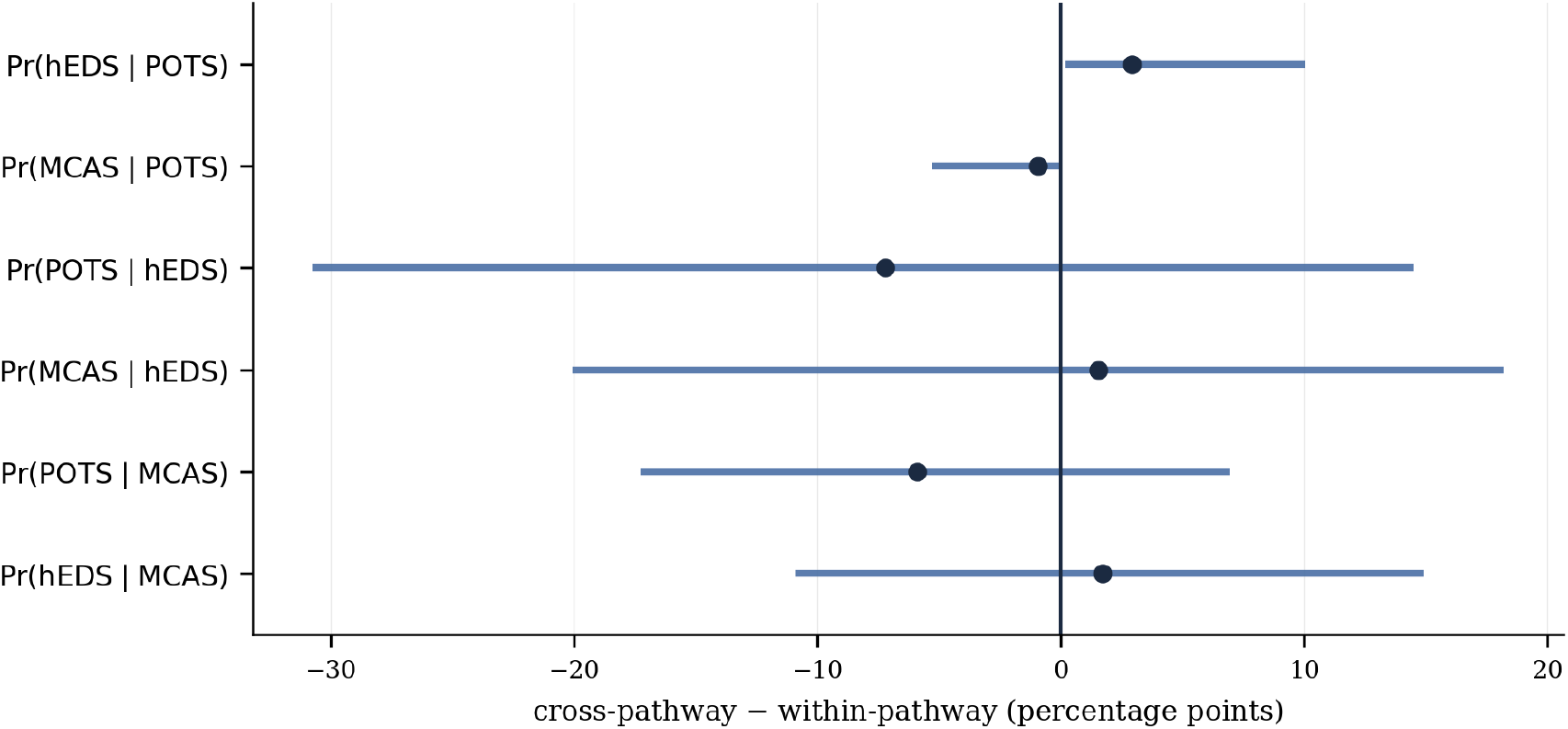
Within-pathway versus cross-pathway symptom evidence, matched at *κ* = 2 symptoms and tempered at 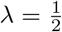. Points are posterior medians of the contrast Δ_*B*|*A*_, bars are 95% credible intervals, and the vertical line marks no difference. The biological prediction—that multi-system (cross-pathway) evidence is uniformly more diagnostic—holds for three of six directions and reverses for the other three. All intervals are plotted in a single colour deliberately: only one excludes zero, and it does so by 0.1 percentage points, so colour-coding it as a positive finding would overstate what Table 4 reports.

The result does not support the biological hypothesis in the strong, uniform form in which it is often stated—and, on the evidence available, does not clearly support it in any form. Two observations must be separated.

First, the *point estimates* exhibit a sign pattern. Cross-pathway evidence raises the posterior relative to within-pathway evidence for ℙ(hEDS | POTS), ℙ(MCAS | POTS) and ℙ(hEDS | MCAS), and lowers it for ℙ(POTS | hEDS), ℙ(MCAS | hEDS) and ℙ(POTS | MCAS). A tempting reading is that the sign tracks whether the target diagnosis is pathway-diffuse: POTS, whose characteristic autonomic symptoms are shared most heavily with the other two conditions, would be predicted *better* by concentrated autonomic evidence, while MCAS and hEDS would be better indicated by evidence spanning systems. We note that even as a description of the point estimates this rule is imperfect: ℙ(MCAS | hEDS) has an MCAS target yet Δ = −5.4 pp, so the rule accounts for five of six directions rather than all six.

Second, and decisively, the *uncertainty* on these contrasts is large. Only one of the six, ℙ(MCAS | POTS) with Δ = +7.6 pp and 95% credible interval [+1.0, +43.9], has an interval excluding zero. The remaining five straddle zero, several by a wide margin. The apparent pattern is therefore not established by these data: it is consistent with them, but so is the null hypothesis of no pathway-dispersion effect in five of six directions. Reporting the sign pattern without the intervals—as an earlier draft of this analysis did—would have manufactured a finding out of Monte Carlo variation in small symptom panels.

We therefore state the conclusion as follows. The pathway decomposition is worth performing, and it generates a specific, biologically motivated, and testable hypothesis: that the value of multisystem symptom evidence depends on the pathway-diffuseness of the target diagnosis. The present corpus does not have the resolution to confirm or refute it. This is a negative result about statistical power, not evidence against the biological account, and the appropriate response is a study designed to test the hypothesis directly rather than a stronger claim from these data.

The contrasts in Table 4 inherit every limitation of Model B, including the unidentified *λ* and the exclusion of incoherent cells, and additionally rest on small per-pathway symptom counts. The signs of Δ are stable, but the magnitudes should not be over-interpreted; the finding is qualitative— that pathway dispersion has a direction-dependent, not uniform, effect—rather than a calibrated effect size.

### 5.4 The monitoring decision

We now answer (Q4) directly. Table 5 reports the threshold-crossing number 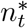 of Definition 4.3.12 for low-(*t* = 0.30) and moderate-(*t* = 0.50) suspicion thresholds, and Figure 6 displays the full accumulation curves against both thresholds.

**Table 5.** Monitoring decision. 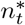 is the minimum number of panel symptoms whose presence raises the posterior median to the threshold *t*; “—” denotes that the four-symptom panel of Section 4.3.2 never reaches *t*. All evaluations at 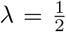. Entries marked *†* are attained at *n* = 0, i.e. the pooled prior already exceeds *t* before any symptom is observed; these carry no symptom information and restate Table 2. Because 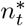 is defined on the posterior median, it ignores the credible interval: for ℙ(hEDS MCAS,) the median crosses 0.30 at one symptom while the interval spans [10.9, 71.4], and the crossing should not be read as a resolved finding.

| Direction | Prior (%) | $n_{0.30}^*$ | $n_{0.50}^*$ |
| --- | --- | --- | --- |
| $\mathbb{P}(\text{hEDS} \mid \text{POTS}, \cdot)$ | 12.1 | 3 | — |
| $\mathbb{P}(\text{MCAS} \mid \text{POTS}, \cdot)$ | 3.9 | — | — |
| $\mathbb{P}(\text{POTS} \mid \text{hEDS}, \cdot)$ | 46.6 | $0^\dagger$ | 4 |
| $\mathbb{P}(\text{MCAS} \mid \text{hEDS}, \cdot)$ | 32.2 | $0^\dagger$ | — |
| $\mathbb{P}(\text{POTS} \mid \text{MCAS}, \cdot)$ | 49.5 | $0^\dagger$ | $0^\dagger$ |
| $\mathbb{P}(\text{hEDS} \mid \text{MCAS}, \cdot)$ | 27.6 | 1 | 3 |

**Figure 6.**
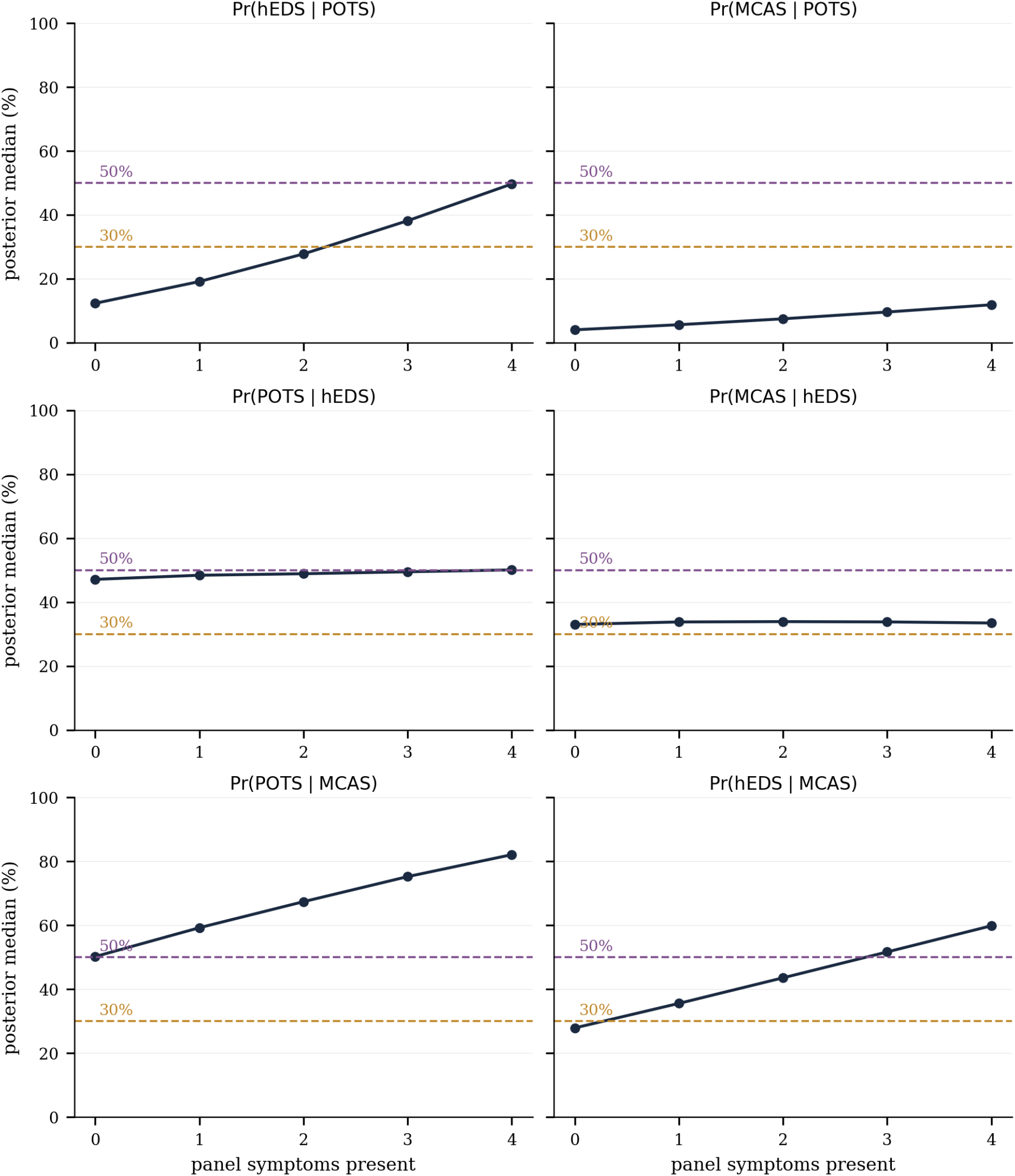
Monitoring-threshold crossing as symptom evidence accumulates, per direction, at 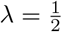. Dashed lines mark the 30% (amber) and 50% (plum) thresholds. The intersection of each curve with a threshold is the corresponding 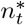 of Table 5.

Three qualitatively distinct regimes emerge, and their clinical readings differ. For directions whose prior already exceeds the low threshold—ℙ(POTS | hEDS), ℙ(MCAS | hEDS), ℙ(POTS | MCAS), all with 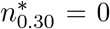—monitoring is warranted on the diagnosis alone, and symptoms serve only to escalate toward the moderate threshold. This furnishes a quantitative basis for the common clinical recommendation to screen hEDS patients for POTS irrespective of presenting symptoms. For ℙ(hEDS | POTS), the low threshold is reached only after three of the four panel symptoms are present, and the moderate threshold is never reached: symptom evidence is *necessary* to justify monitoring, and a symptom-sparse POTS patient does not meet even the low-suspicion bar for hEDS. Most starkly, ℙ(MCAS | POTS) never reaches even the low threshold within the panel: on this evidence base, no symptom profile short of direct mast-cell testing raises MCAS suspicion in a POTS patient to an actionable level—a finding that concords with the low pooled prior and the wide uncertainty of that direction, and that argues *against* routine symptom-driven MCAS monitoring in POTS absent other indications.

Definition 4.3.12 deliberately stops short of a cost-calibrated optimal rule. The threshold *t* encodes a cost ratio between missed comorbidity and unnecessary investigation that this evidence base cannot supply. The layer therefore answers the estimable half of (Q4)—how much symptom evidence is required to reach a stated suspicion level—while leaving the normative choice of that level explicit and external. This is, we argue, the appropriate division of labour between a statistical analysis and a clinical guideline.

### 5.5 Model C: latent structure and its obstruction

#### 5.5.1 Non-identifiability, empirically demonstrated

Corollary 4.3.15 asserts that any latent class fit to marginal data is determined by initialization rather than evidence. We verify this directly. The eight-patient supplement admits the joint symptom patterns required to *run* (17)–(18), but at *N* = 8 with *J* = 13 symptoms the model carries (*K* − 1) + *KJ* = 27 free parameters for *K* = 2, a parameter-to-observation ratio of 3.4. Executing the validated estimator (below) on these eight patients from twelve random initializations yields a smaller-class proportion ranging over [0.249, 0.500]—the same data returning materially different “structure” according to the seed. This is the empirical signature of Corollary 4.3.15: convergence is achieved every time, and every time it is meaningless.

#### 5.5.2 Estimator validation on synthetic data

To establish that the obstruction lies in the data and not the code, we applied the same EM implementation to synthetic patient-level data generated from a known two-class structure (*N* = 1200, *J* = 8, ***π*** = (0.6, 0.4)). BIC selected 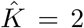 correctly (Figure 7), and the recovered class proportions (0.382, 0.618) matched the generative (0.40, 0.60) to within Monte Carlo error. The estimator is therefore correct; the patient-level corpus is simply too small and, more fundamentally, the aggregate corpus is non-identified.

**Figure 7.**
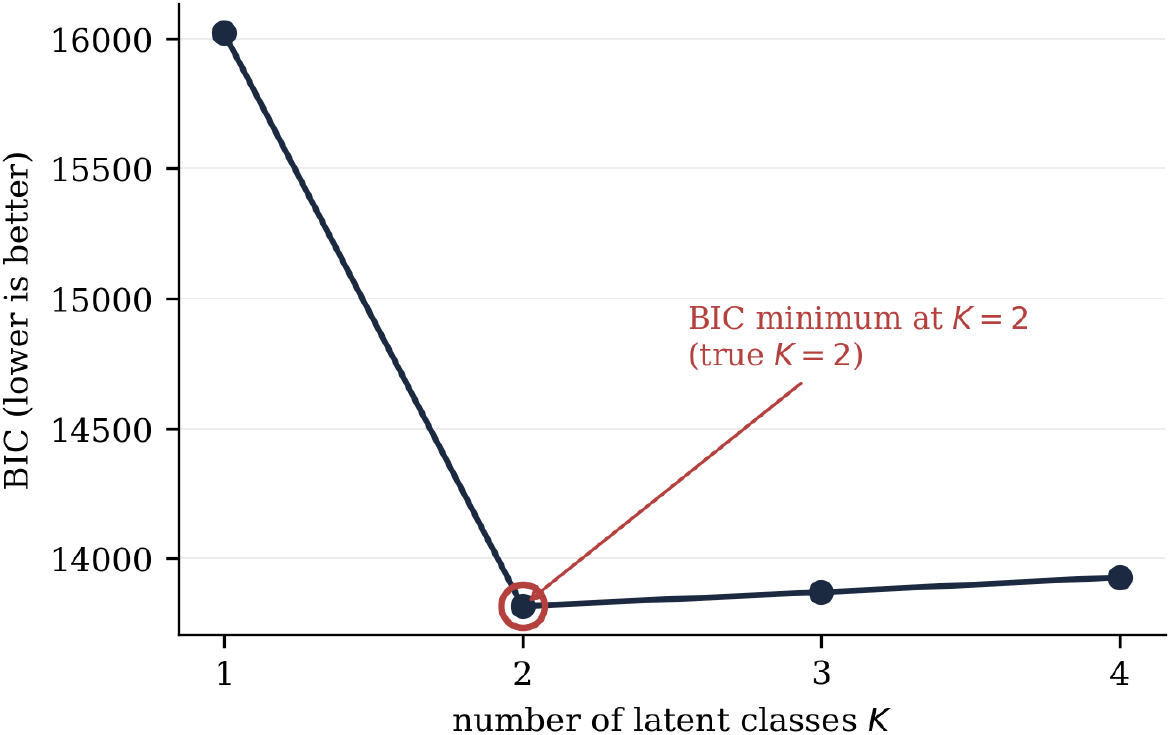
Validation of the latent-class EM estimator on synthetic data with known *K* = 2. BIC is minimized at 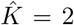, and class proportions are recovered accurately. This certifies the implementation, isolating the failure on real data as a property of the data rather than the method.

#### 5.5.3 Power analysis: the binding constraint is ascertainment

A simulation sweeping sample size and ascertainment completeness (Table 6) shows that recovery of a known two-class structure requires on the order of 10^2^ patients under complete ascertainment, but *fails at every sample size tested* when half the symptom entries are unascertained and coded as absent—the coding regime forced by the “unclear” entries pervading the patient-level supplement, where 52% of cells are unascertained. The operational implication is that ascertainment completeness, not patient count, is the design parameter that governs feasibility of a future latent-class study.

**Table 6.** Simulation-based recovery rate of a known *K* = 2 structure (fraction of replicates in which BIC (Nylund et al., 2007) selects 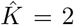), by sample size and ascertainment regime. “Complete” denotes fully observed indicators; “50% unascertained” codes half the entries as absent, mimicking the supplement’s “unclear” entries.

| Regime | $N=25$ | $N=50$ | $N=100$ | $N=200$ | $N=400$ | $N=800$ |
| --- | --- | --- | --- | --- | --- | --- |
| Well-separated, complete | 100 | 100 | 100 | 100 | 100 | 100 |
| Moderate sep., complete | 12 | 32 | 68 | 100 | 100 | 100 |
| Moderate sep., 50% unasc. | 0 | 0 | 0 | 0 | 0 | 20 |
| Weak sep., 50% unasc. | 0 | 0 | 0 | 0 | 0 | 0 |

#### 5.5.4 A defensible aggregate substitute: profile correlation

Since latent structure is unavailable, we report the estimable weaker object: the correlation between the three diseases’ pooled symptom-prevalence profiles across the 14 symptoms observed in all three. With posterior propagation, the POTS–hEDS profile correlation is *r* = 0.624 (95% CrI [0.358, 0.815]), an interval excluding zero; the two correlations involving MCAS have intervals straddling zero (POTS–MCAS: 0.325, [−0.073, 0.584]; hEDS–MCAS: 0.241, [−0.187, 0.550]). Thus the only defensible similarity claim is between POTS and hEDS, consistent with the Model A finding that this pair is the best-evidenced axis of the triad. Figure 8 displays the profiles ordered by cross-disease differentiation.

**Figure 8.**
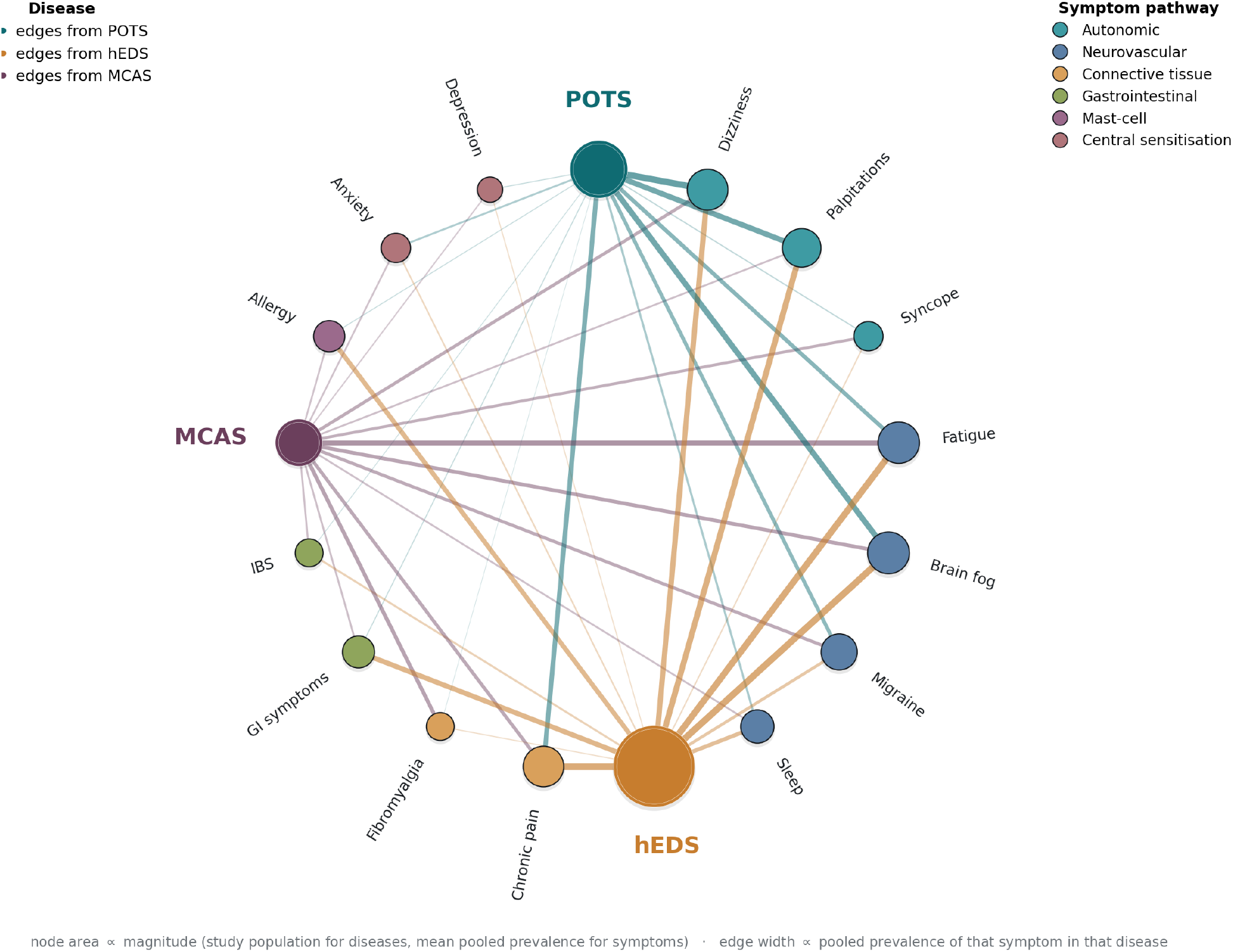
Shared-symptom network across the three diagnoses, drawn in the convention of a network meta-analysis diagram: all nodes lie on a single circle, node area is proportional to magnitude (study population for diagnoses, mean pooled prevalence for symptoms), and edge width is proportional to the pooled prevalence of that symptom in that diagnosis. Symptoms are coloured by biological pathway and placed in contiguous arcs, with each diagnosis positioned adjacent to the pathway it characterises. Symptom names are abbreviated to their leading term; full symptom definitions are as given in the data description. The figure is the graphical counterpart of the profile-similarity analysis, whose only interval-excludes-zero correlation is POTS–hEDS.

### 5.6 The single recoverable joint table

Proposition 4.3.19 applies to exactly one cohort in the corpus: the MCAS-selected retrospective case series of Weinstock et al. (2023), comprising *N* = 8 patients, all MCAS-diagnosed, of whom four were diagnosed with POTS, four with hEDS, and two with both. Since the cohort is MCAS-selected, “all three” coincides with {POTS, hEDS} within it, and (19) yields

|  | hEDS<sup>+</sup> | hEDS<sup>−</sup> | total |
| --- | --- | --- | --- |
| POTS<sup>+</sup> | 2 | 2 | 4 |
| POTS<sup>−</sup> | 2 | 2 | 4 |
| total | 4 | 4 | 8 |

The table is exactly balanced. Under independence one would expect 8*×*0.5*×*0.5 = 2 doubly-affected patients; two are observed. The conditional odds ratio is therefore

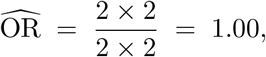

with Woolf standard error (1955) 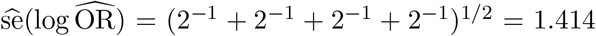, giving a 95% interval exp(log 1.00 *±* 1.96 *×* 1.414) = [0.063, 15.99]. Fisher’s exact test (Fisher, 1922) returns *p* = 1.000. The interval spans more than two orders of magnitude, and the point estimate sits exactly at the null.

#### Remark 5.6.1

(What the joint table does and does not establish). This is a null result, and it is important to characterise it correctly. It is *not* evidence that POTS and hEDS are independent within

MCAS populations; with four patients in each margin the test has essentially no power, and the credible interval is consistent with anything from a strong negative to a strong positive association. What the table does establish is narrower and, for the present paper, more consequential: the corpus contains *no* usable individual-level joint information about the triad. The one cohort that reports the trivariate structure required by Proposition 4.3.19 is too small to constrain the joint distribution at all.

#### Remark 5.6.2

(Correction of an earlier analysis). An earlier version of this analysis recorded this cohort with *N* = 139 and marginals (25.2%, 23.7%, 15.1%), from which it derived 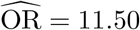 with *p* = 4.5 *×* 10^−8^, and reported that estimates of triad prevalence constructed under independence understate the truth by a factor of 2.53. On verification against the published source those figures proved to be an extraction error: the value 139 appears in Weinstock et al. (2023) only within a citation to an unrelated mastocytosis cohort, and the percentages do not correspond to any quantity reported there. The corrected extraction, given above and cross-checked against the paper’s own summary statements (“POTS in four”, “Four subjects had hEDS”), reverses the finding entirely: from a highly significant association to an exact null. We record this explicitly because the erroneous result was the strongest single claim in the earlier analysis, and because its correction illustrates the paper’s broader thesis with uncomfortable directness—an apparently decisive individual-level finding, on inspection, was an artifact of the aggregation pipeline rather than a property of the evidence.

## 6 Discussion

### 6.1 Which claims the evidence supports

The analysis licenses a graded set of conclusions. Most secure are the *directed comorbidity probabilities* of Model A, understood as within-referred-population quantities with honestly wide intervals; reassuringly, the pooled ℙ(hEDS | POTS) = 12.1% coincides closely with the 13% reported independently by Yao et al. (2025) under strict criteria. Next is the *conditional association* between POTS and hEDS: both the Model C profile correlation (*r* = 0.62, excluding zero) is the only positive evidence for it, since the recovered 2 *×* 2 table proved on correction to be an exact null (OR = 1.00, *p* = 1.000) with essentially no power. The POTS–hEDS association therefore rests on a single aggregate-level analysis and should be regarded as suggestive rather than corroborated. The *pathway-dispersion* contrast of Section 5.3 is, by contrast, not supported: five of its six credible intervals include zero, and the apparent sign pattern—though biologically interpretable—cannot be distinguished from Monte Carlo variation in small symptom panels. We retain it as a stated hypothesis for future testing and explicitly decline to claim it as a result. The *monitoring thresholds* of Section 5.4 are supported as conditional statements: given the pooled priors and the tempered update, they identify which directions warrant monitoring on diagnosis alone (hEDS → POTS, hEDS → MCAS, MCAS → POTS) and which require substantial symptom evidence (POTS → hEDS) or cannot reach an actionable level at all (POTS → MCAS). Least secure, indeed unsupported, are any *latent-phenotype* or *causal* claims: Theorem 4.3.14 and Proposition 4.3.18 show these are not merely unproven but non-identifiable from the available designs.

The pooled ℙ(MCAS | POTS) = 4.0% warrants separate comment, since it sits far below the 37% that Yao et al. (2025) report for MCAS under consensus-2 criteria. This order-of-magnitude discrepancy is not noise but a direct manifestation of the definitional heterogeneity discussed below: consensus-1 and consensus-2 MCAS criteria differ by roughly an order of magnitude in ascertained prevalence, and the corpus pools across both. The discrepancy is thus the clearest empirical instance of the coherence problem the paper formalizes, and it cautions against reading any single MCAS-directed estimate without reference to the criteria that generated it.

### 6.2 Collider bias as the principal alternative explanation

Any association recovered from this corpus—including the POTS–hEDS profile correlation of Section 5.5, and any that a larger trivariate cohort might yield in future—demands scrutiny before it is read as biological clustering. Every cohort in the corpus is assembled at a specialty clinic on the basis of already carrying one of the three diagnoses. Conditioning on referral is conditioning on a collider: if POTS and hEDS each independently raise the probability of specialist referral, then within a referred sample they will appear negatively or positively associated even if independent in the source population, by the mechanism of Berkson’s paradox (1946). The direction and magnitude of this bias depend on the referral probabilities, which are unobserved. Thus any observed association is consistent with (i) genuine shared aetiology, (ii) Berksonian selection (Griffith et al., 2020; Hernán et al., 2004), or (iii) a mixture; the data cannot apportion between them. We regard this as the single most important limitation on the clinical interpretation of the results, and note that it applies with equal force to the wider triad literature, much of which reports referral-clinic associations as though they estimated population parameters. The point is sharpened, not weakened, by the null of Section 5.6: the only individual-level table available is uninformative, so the collider question cannot even be posed empirically here.

### 6.3 The coherence finding and the myth of a single population

The two outright failures of the feasibility constraint (13), and the partial infeasibility detected in most remaining cells, are on reflection unsurprising given that each pooled marginal is assembled from a different set of studies applying different diagnostic criteria—most consequentially the competing “consensus-1” and “consensus-2” definitions of MCAS (Valent et al., 2012; Afrin et al., 2021), which differ in stringency by an order of magnitude in reported prevalence. When marginals estimated under incompatible definitions are combined, no single joint law need exist, and the incoherence fraction detects exactly this. The methodological moral is that meta-analytic pooling of prevalences across a definitionally fractured literature can produce numbers that are individually defensible yet jointly impossible. We emphasize that the diagnostic identified few hard failures here; its contribution is to make the possibility checkable at all, and a corpus with more overlapping symptom reporting would submit to a far sharper test.

### 6.4 Statistical significance versus practical resolving power

Model B exhibits the familiar divergence between significance and utility. Individual symptoms carry likelihood ratios whose credible intervals frequently include unity (Figure 3); only in combination do they move the posterior substantially, and then only under an independence assumption that inflates their joint evidential value. A predictor whose apparent discrimination depends on multiplying correlated, weakly-informative, and partly incoherent inputs is not a clinically deployable instrument. The tempered curve of Figure 4 should be read less as a calibrated prediction than as a demonstration of how much apparent confidence is manufactured by the independence assumption.

### 6.5 Summary of estimated comorbidity probabilities

The preceding sections express results as credible intervals, likelihood ratios and tempered posteriors. It is useful to consolidate them into the single quantity of clinical interest: for a patient already carrying one diagnosis, the estimated probability of carrying a second, as a function of how many of the relevant symptoms are present. Table 7 reports this. All figures are posterior medians under the tempered update 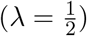, rounded to whole percentages.

**Table 7.** Estimated probability of a second diagnosis, given an existing diagnosis and the number of relevant symptoms present. A row reads: a patient who already has the first condition, and who presents this many of the four *panel* symptoms listed below for that row, has approximately this probability of also having the second condition. The panel is the four most informative of the four to ten symptoms retained for that direction (Section 4.3.2); “all four” therefore means all four panel symptoms, not all symptoms recorded in the corpus. Values are rounded posterior medians at 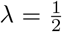; the associated uncertainty is substantial and is reported in Tables 2 and 3.

| Existing diagnosis | Probability of also having | No symptoms | Three symptoms | All four symptoms |
| --- | --- | --- | --- | --- |
| POTS | hEDS | 12% | 28% | 50% |
| POTS | MCAS | 4% | 7% | 12% |
| hEDS | POTS | 47% | 49% | 50% |
| hEDS | MCAS | 33% | 34% | 33% |
| MCAS | POTS | 50% | 67% | 82% |
| MCAS | hEDS | 28% | 44% | 60% |
| bottomrule |  |  |  |  |

The four panel symptoms differ by row, since the symptoms that carry information about one diagnosis are not those that carry information about another. They are:

- **POTS** → **hEDS:** allergy symptoms, GI symptoms, IBS, fatigue, chronic pain, syncope.
- **POTS** → **MCAS:** fibromyalgia, allergy symptoms, fatigue, migraine, anxiety, brain fog.
- **hEDS** → **POTS:** anxiety, migraine, dizziness, palpitations, syncope, chronic pain.
- **hEDS** → **MCAS:** constipation, depression, anxiety, migraine, abdominal pain, fatigue.
- **MCAS** → **POTS:** presyncope, dizziness, flushing, brain fog, anxiety, migraine.
- **MCAS** → **hEDS:** allergy symptoms, sleep disturbance, dizziness, fatigue, abdominal pain, migraine.

First, these are *group-level* quantities: they describe how frequently the second diagnosis occurs among patients with that profile in the published cohorts, and do not constitute individual risk predictions. Second, the associated uncertainty is large—for several rows the credible interval spans thirty percentage points or more—so differences of a few points between cells carry no interpretive weight. Third, every contributing cohort was recruited at a specialist clinic, so these figures describe patients already referred for investigation and will overstate the corresponding probabilities in an unselected population.

#### Remark 6.5.1

(The absence of a symptoms-only estimate). Table 7 is necessarily conditioned on an existing diagnosis. The corresponding quantity for a patient with *no* established diagnosis is not estimable from this evidence base: every study in the corpus recruited patients already carrying one of the three conditions, so the data are uninformative about the prevalence of these diagnoses in an unselected population. Producing such an estimate would require an external general-population prevalence prior, and for MCAS no reliable estimate exists (Section 7).

## 7 Limitations

We collect the principal threats to validity, several already noted in situ.

- **Referral and collider bias**. All estimands are conditioned on specialty referral; none estimates a general-population parameter. This applies to every estimate reported here (Section 6).
- **Definitional heterogeneity**. Competing MCAS criteria, and evolving hEDS and POTS definitions, mean the pooled marginals may not describe a common population; the coherence diagnostic quantifies where this becomes self-contradictory.
- **Small** *k*. Three of six directions rest on *k* ≤ 3 studies and two on *k* = 2; the wide prediction intervals are appropriate but limit inferential power. Publication-bias diagnostics (funnel plots and Egger’s test (1997)) are uninformative at these *k* and were deliberately not computed, in line with standard guidance against their use for *k <* 10 (Sterne et al., 2011).
- **Assumption 4.3.3**. Model B’s identification of the reference group rests on ℙ(*S* | *A, B*) ≈ ℙ(*S* | *B*), which is unverifiable in this corpus and whose failure is entangled with definitional incoherence.
- **Unidentified dependence parameter** *λ*. The tempered update introduces *λ* as a sensitivity parameter that no data can calibrate; results are reported as intervals over 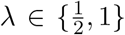 rather than as point predictions.
- **Informative missingness**. MCAS is disproportionately unreported in POTS- and hEDS-selected cohorts, a missing-not-at-random (Little & Rubin, 2019; Rubin, 1976) pattern that biases the directions into MCAS by an unknown amount.
- **No usable joint information**. The only cohort reporting the trivariate structure required by Proposition 4.3.19 contains eight patients, and the recovered table is exactly balanced (OR = 1.00, *p* = 1.000). The corpus therefore constrains the joint distribution of the three diagnoses not at all. An earlier extraction of this cohort was erroneous and produced a spurious strong association; the correction is documented in Remark 5.6.2.
- **Latent structure genuinely inaccessible**. By Theorem 4.3.14 and Proposition 4.3.18, the latent-phenotype question cannot be answered by any meta-analysis of disease-selected cohorts, a limitation of design rather than of sample size.
- **Panel truncation**. The evaluation panel is capped at six symptoms per direction for tractability, excluding three to four symptoms that passed the coherence screen (Section 4.3.2). Reported probabilities are therefore conditioned on a subset of the usable evidence, and the phrase “all four symptoms” throughout refers to the panel rather than to the full symptom record.
- **Pathway contrasts rest on thin panels**. The pathway-dispersion analysis draws on small per-pathway symptom counts, so the signs of Δ_*B*|*A*_ are more trustworthy than their magnitudes; the finding is qualitative and inherits Model B’s assumptions in full.
- **Monitoring thresholds are not cost-calibrated**. The decision layer reports how much symptom evidence reaches a stated threshold *t*, but *t* encodes an unestimated cost ratio between missed comorbidity and unnecessary investigation; the normative choice of *t* is left external, and the thresholds should not be read as an optimized screening policy.
- **Disease-specific symptoms unrepresented**. Generalized joint hypermobility, the one putatively hEDS-specific manifestation, is absent from the poolable corpus, so Models A and B quantify the evidential content of convergent symptoms only.

### 7.1 Ascertainment by self-report

The pooled estimates are dominated by self-selected surveys in which the comorbidity being estimated is itself self-reported. Shaw et al. (2019) supplies 88% and 89% of the pooled sample for the two POTS-cohort estimands; Daylor et al. (2025) and Wachs & Papendorp (2026) together supply 94% and 96% for the two hEDS-cohort estimands; Weinstock et al. (2025) supplies 99% of both MCAS-cohort estimands. Random-effects shrinkage moderates the influence of large studies but does not reverse it.

This matters beyond the usual caution about survey data. A respondent who has encountered the hypothesis of a triad is more likely to report all three diagnoses, so the association under study can be manufactured by differential awareness among respondents. This is a distinct mechanism from the collider bias of Section 6.2: it does not require selection on a common effect, only correlated reporting. It is not detectable within the corpus, and we cannot bound it. Only Kozyra et al. (2024), a claims cohort with physician-coded diagnoses, has ascertainment independent of patient report, and it contributes three symptom prevalences and no comorbidity marginal. A replication restricted to clinician-ascertained cohorts is the single most valuable next study.

### 7.2 An unmodelled common cause

Hereditary alpha-tryptasemia associates with both joint hypermobility and mast-cell mediator symptoms, and is present in a non-trivial fraction of the general population. It is therefore a candidate common cause of part of the hEDS–MCAS association estimated here. No study in the corpus reports tryptase genotype, so we cannot adjust for it, and *π*_MCAS|hEDS_ should be read as containing an unknown contribution from this source.

### 7.3 Reproducibility of the pathway partition

The six pathways of Section 4.3.2 are those established in the biological background of Sections 2.2–2.7. The assignment of individual symptoms to them, however, was made for the present analysis: it is not empirically derived, was not pre-registered, and the pathway contrast is sensitive to it. The full mapping is given in Table A3. Its central premise—that each symptom belongs to exactly one pathway—is a simplification that our own background section contradicts in at least three places: fatigue is there attributed to autonomic, neurovascular and central-sensitisation mechanisms alike, and brain fog and sleep disturbance to two apiece. Such symptoms were assigned to a single most proximate pathway. Readers who would assign them differently should expect the contrasts of Table 4 to move, and this is a further reason to treat that analysis as exploratory.

## 8 Future directions

Three developments would materially advance the question. First, an *unselected* population sample—or a cohort selected on a variable other than the three diagnoses—would break the collider structure of Proposition 4.3.18 and render the trivariate table, and hence latent structure, identifiable. Second, the power analysis of Table 6 specifies a concrete design target: on the order of 200 patients with *near-complete* symptom ascertainment (≳ 80%) would suffice to identify a two-class structure, whereas larger but sparsely-ascertained samples would not; ascertainment protocol is the binding constraint. Third, individual-level data would permit direct estimation of the symptom dependence structure, replacing the unidentified *λ* of (15) with an estimated quantity and allowing the conditional-independence assumption (3) to be tested rather than assumed. Each of these is a data-collection prescription; none can be met by further modelling of the existing aggregate corpus, whose informational ceiling the present analysis has, we believe, essentially reached.

## 9 Conclusion

We posed four questions: how strongly the three conditions co-occur, whether symptoms sharpen the prediction of a second diagnosis given a first, whether the triad reflects a shared latent phenotype, and whether a symptom profile can be turned into a monitoring decision. To the first we give quantitative if wide-intervalled answers, with the caveat that all are referral-conditioned. To the second we give a qualified yes under an independence assumption we show to be false, and a more sober assessment—an interval bounded below by the tempered update and above by the naive one—once that assumption is relaxed; the practical resolving power of symptoms is modest, and, as the pathway-structured analysis shows, not resolvable at this sample size: the contrast between multi-system and single-system symptom evidence has a credible interval excluding zero in only one of six directions, so the biologically motivated hypothesis it encodes remains open. To the third we give a principled negative: latent structure is non-identifiable from the available designs, a fact we establish as a theorem rather than lament as a data shortage.

To the fourth we give the paper’s most directly clinical answer. The decision layer converts the posteriors into a monitoring rule and yields a graded, actionable triage: hEDS patients warrant POTS and MCAS surveillance on diagnosis alone; MCAS patients warrant POTS surveillance likewise; a POTS patient warrants hEDS surveillance only once symptom evidence accumulates (three of four panel symptoms for low suspicion); and, most usefully as a *negative* guide, no symptom profile in the corpus raises MCAS suspicion in a POTS patient to an actionable level, so symptom-driven MCAS screening in POTS is not supported and direct testing is the only informative route. These are exactly the “potential and limitations” the investigation set out to weigh: the symptom data are genuinely useful for some monitoring decisions and genuinely uninformative for others, and the analysis says which is which.

The two methodological contributions we regard as transportable beyond this application are the *coherence diagnostic*, which turns the Fréchet feasibility inequalities into an audit of whether separately-pooled marginals can describe a common population, and the explicit *identifiability analysis* of latent structure under disease-selected sampling. Both express a single discipline: distinguishing what the data can determine from what one might wish them to determine. The recurring lesson of the analysis is that the triad literature’s confident causal and phenotypic claims outrun their evidential base—but that, held to a correct accounting of uncertainty, the same data still yield a defensible and clinically legible set of monitoring recommendations. That accounting—wide intervals, excluded incoherent cells, an unidentified dependence parameter, a non-identifiability theorem, and thresholds that some directions simply cannot reach—is not a failure of the analysis but the precondition for trusting the decisions it does support.

## Data Availability

The data analysed in this study were obtained exclusively from previously published, publicly available scientific articles identified through the systematic literature search described in the manuscript. All source studies are cited in the References section. Derived data generated during the analyses is either contained within the manuscript or can be asked for upon reasonable request to the authors

## Declarations

| Item | Statement |
| --- | --- |
| Ethical approval | This study analysed only aggregate data from previously published studies. No human participants were recruited and no individual-level identifiable data were accessed. Ethical approval was therefore not required. |
| Data availability | All data analysed here are aggregate figures drawn from the published studies listed in Table A1. The study-level inputs, the variables extracted from each source, and the symptom-to-pathway partition are reported in full in Appendix Tables A1–A3; no data beyond those tables were used. |
| Article type | Original research: a quantitative evidence synthesis presenting new pooled estimates, a new feasibility diagnostic for meta-analytic marginals, and an identifiability result. It is not a review article. |
| Reporting standard | No protocol was registered and the literature search was not exhaustive, so PRISMA reporting items are not claimed and the synthesis should not be read as a systematic review. Eligibility criteria, the full study inventory and the variables extracted from each source are reported in Section 3.2 and Appendix Tables A1–A3. |
| Funding | None. |
| Competing interests | The authors declare no competing interests. |
| Author contributions | Both authors contributed to study design, data extraction, analysis and manuscript preparation, and approved the final version. |

## Appendix Source Data and Database Construction

This appendix documents the provenance of every quantity entering the analysis. No primary human-subjects data were collected for this work: the corpus is assembled entirely from prevalence figures reported in the published literature. Table A1 lists the source studies and Table A2 the variables extracted from them.

Twenty-two rows are curated in total, all of which have been traced to an identifiable published source. Two required attribution during preparation of this appendix: the row entered without a study identifier is the claims-database cohort of Kozyra et al. (2024), and the row labelled by its selection symptom is the Houston Methodist series of Quigley et al. (2024). Nineteen rows enter the primary analysis and three are restricted to the sensitivity variant.

**Table A1.** Source studies. *Primary cohort* is the diagnosis on which the study sample was selected. *Data extracted* records the number of symptom-prevalence fields populated and any non-degenerate diagnostic marginals; a cohort’s own diagnosis is excluded, being identically 100% and therefore uninformative by Proposition 4.3.18. Rows marked *†* carry non-canonical cohort labels and enter only the sensitivity variant.

| Study | Year | Primary cohort | $N$ | Data extracted |
| --- | --- | --- | --- | --- |
| Kohno et al. | 2021 | POTS | 69 | 11 symptom prevalence |
| Afrin et al. | 2017 | MCAS | 413 | 20 symptom prevalences |
| Rombaut et al. | 2010 | hEDS | 32 | 3 symptom prevalences |
| Bénistan & Martinez | 2019 | hEDS | 37 | 7 symptom prevalences |
| Wachs & Papendorp | 2026 | hEDS | 2,572 | 8 symptom prevalences; comorbidity: POTS, MCAS |
| Deb et al. | 2015 | POTS | 39 | 12 symptom prevalences; comorbidity: hEDS |
| Daylor et al. | 2025 | hEDS | 3,360 | 12 symptom prevalences; comorbidity: POTS, MCAS |
| Shaw et al. | 2019 | POTS | 4,835 | 13 symptom prevalences; comorbidity: hEDS, MCAS |
| Weinstock et al. | 2025 | MCAS | 553 | 15 symptom prevalences; comorbidity: POTS, hEDS |
| De Wandele et al. | 2014 | hEDS | 39 | 3 symptom prevalences; comorbidity: POTS |
| Angeli et al. | 2024 | POTS | 378 | 10 symptom prevalences |
| Lam et al. | 2021 | hEDS | 603 | 12 symptom prevalences |
| Weinstock et al. | 2023 | MCAS | 8 | 1 symptom prevalence; comorbidity: POTS, hEDS, triad |
| Jennings et al. | 2014 | MCAS | 420 | 18 symptom prevalences |
| Kozyra et al. | 2024 | hEDS | 19,790 | 3 symptom prevalences |
| Celletti et al. | 2020 | hEDS | 102 | 1 symptom prevalence; comorbidity: POTS |
| Novak et al. | 2025 | hEDS | 270 | 6 symptom prevalences; comorbidity: POTS, MCAS |
| Thwaites et al. | 2022 | hEDS | 45 | 2 symptom prevalences |
| van Middendorp et al. | 2025 | POTS | 599 | 4 symptom prevalences; comorbidity: hEDS, MCAS |
| Bayrak et al.$^\dagger$ | 2024 | POTS (paediatric) | 28 | 11 symptom prevalence; comorbidity: POTS |
| Song et al.$^\dagger$ | 2021 | Mixed EDS (76% hEDS) | 98 | 8 symptom prevalences; comorbidity: hEDS, MCAS |
| Quigley et al.$^\dagger$ | 2024 | Nausea/vomiting | 26 | 4 symptom prevalences; comorbidity: POTS, hEDS, MCAS |

**Table A2.** Variables extracted. All symptom and diagnostic fields are study-level prevalences reported as percentages of the study cohort; counts were reconstructed via Definition 4.2.2. No individual-level records were used except in the patient-level supplement of Section 4.2.

| Variable | Source | Type | Studies |
| --- | --- | --- | --- |
| dizziness / orthostatic intolerance / dysautonomia | Study prevalence | Percentage | 12 |
| fatigue | Study prevalence | Percentage | 11 |
| palpitations / tachycardia / cardiac symptoms in general | Study prevalence | Percentage | 8 |
| syncope | Study prevalence | Percentage | 8 |
| presyncope | Study prevalence | Percentage | 4 |
| migraine / headache | Study prevalence | Percentage | 15 |
| anxiety | Study prevalence | Percentage | 12 |
| depression | Study prevalence | Percentage | 11 |
| allergy symptoms | Study prevalence | Percentage | 7 |
| skin rash / itching | Study prevalence | Percentage | 4 |
| GI symptoms | Study prevalence | Percentage | 12 |
| Hives / urticaria | Study prevalence | Percentage | 4 |
| Sleep | Study prevalence | Percentage | 5 |
| Brain fog | Study prevalence | Percentage | 7 |
| Flushing | Study prevalence | Percentage | 5 |
| Tremor | Study prevalence | Percentage | 5 |
| IBS | Study prevalence | Percentage | 8 |
| Fibromyalgia | Study prevalence | Percentage | 6 |
| Abdominal pain | Study prevalence | Percentage | 6 |
| Nausea / vomiting | Study prevalence | Percentage | 5 |
| Anaphylaxis | Study prevalence | Percentage | 1 |
| GERD / reflux | Study prevalence | Percentage | 5 |
| Hypotension | Study prevalence | Percentage | 1 |
| Constipation / diarrhea | Study prevalence | Percentage | 6 |
| dysphagia | Study prevalence | Percentage | 1 |
| Bloating / dyspepsia | Study prevalence | Percentage | 2 |
| cardiac disease | Study prevalence | Percentage | 2 |
| Muscle weakness | Study prevalence | Percentage | 3 |
| chronic pain / fibromyalgia | Study prevalence | Percentage | 8 |
| POTS diagnosis | Cohort marginal | Percentage | 14 |
| hEDS diagnosis | Cohort marginal | Percentage | 17 |
| MCAS diagnosis | Cohort marginal | Percentage | 11 |
| All three diagnoses | Cohort marginal | Percentage | 1 |
| Cohort type | Study design | Categorical | 22 |
| Sample size $n_i$ | Study report | Count | 22 |

**Table A3.** Symptom-to-pathway partition used in Section 4.3.2. The six pathways are those established in Sections 2.2–2.7; the assignment of individual symptoms to them was made for the present analysis and is neither empirically derived nor pre-registered. It is stated in full so that it can be checked and disputed. Three assignments are contestable and are marked *‡*: Sections 2.3, 2.6 and 2.7 each describe fatigue as arising through their respective mechanisms, and brain fog and sleep disturbance are each described in two. These were assigned to the single most proximate pathway, which is a modelling simplification rather than a biological claim, and the contrast of Table 4 is sensitive to it.

| Pathway | Section | Symptoms |
| --- | --- | --- |
| Connective tissue | 2.2 | muscle weakness |
| Autonomic | 2.3 | cardiac disease<sup>‡</sup>, dizziness / orthostatic intolerance, hypotension, palpitations / tachycardia, presyncope, syncope, tremor |
| Mast-cell | 2.4 | allergy symptoms, anaphylaxis, flushing, hives / urticaria, skin rash / itching |
| Gastrointestinal | 2.5 | abdominal pain, bloating / dyspepsia, constipation / diarrhoea, dysphagia, GERD / reflux, GI symptoms, IBS, nausea / vomiting |
| Neurovascular | 2.6 | brain fog<sup>‡</sup>, migraine / headache |
| Central sensitisation | 2.7 | anxiety, chronic pain / fibromyalgia, depression, fatigue<sup>‡</sup>, fibromyalgia, sleep<sup>‡</sup> |

